# Evaluation of Guillain-Barré syndrome following Respiratory Syncytial Virus Vaccination among Medicare Beneficiaries 65 Years and Older

**DOI:** 10.1101/2024.12.27.24319702

**Authors:** Patricia C. Lloyd, Purva B. Shah, Henry T. Zhang, Nimesh Shah, Narayan Nair, Zhiruo Wan, Mao Hu, Tainya C. Clarke, Meng Chen, Xinxin Lin, Rose Do, Jing Wang, Yue Wu, Yoganand Chillarige, Richard A. Forshee, Steven A. Anderson

## Abstract

**Importance:** The United States Food and Drug Administration conducts routine post-market surveillance of approved vaccines to identify and characterize health outcomes risk associated with vaccination. Pre-licensure clinical trials of RSVPreF3+AS01 and RSVPreF vaccines identified a small number of Guillain-Barré syndrome cases, a serious acute demyelinating disease, following vaccination.

**Objective:** To use Centers for Medicare & Medicaid Services Medicare Fee-for-Service administrative claims and Medicare Part D data to evaluate risk of Guillain-Barré syndrome following respiratory syncytial virus vaccination.

**Design, Settings, and Participants:** We studied vaccines administered between May 3, 2023, when vaccines were first approved, through January 28, 2024. This self-controlled case series study design compared the incidence of Guillain-Barré syndrome during risk intervals of 1–42 days following vaccination to the incidence in subsequent control intervals (43–90 days) among Medicare beneficiaries, aged 65 years and older, enrolled in Fee-for-Service and Medicare Part D.

**Exposures:** Receipt of a single dose of RSVPreF3+AS01 or RSVPreF vaccines.

**Outcomes:** Guillain-Barré syndrome was identified using diagnosis code G61.0 in the primary diagnosis position on hospital inpatient claims; returned cases were confirmed via medical record review using Brighton Collaboration’s case definition.

**Results:** We captured approximately 3.23 million Medicare beneficiaries that received respiratory syncytial virus vaccination during the study period. A total of 95 incident Guillain-Barré syndrome cases were observed following respiratory syncytial virus vaccination. In our chart-confirmed self-controlled case series analysis, we observed an increased incidence of Guillain-Barré syndrome following RSVPreF3+AS01 (IRR: 2.46, [95% CI: 1.19, 5.08]) with an attributable risk of 6.5 cases per 1 million doses. We observed an increased, yet not statistically significant, incidence of Guillain-Barré syndrome following RSVPreF (IRR: 2.02, [95% CI: 0.93, 4.40]) with an attributable risk of 9 cases per 1 million doses.

**Conclusion and Relevance:** Findings from this self-controlled case series study suggest an increased risk of Guillain-Barré syndrome during 1-42 days following respiratory syncytial virus vaccination, with fewer than 10 excess Guillain-Barré syndrome cases per 1 million vaccine doses among Medicare beneficiaries 65 years and older. The U.S. Food and Drug Administration maintains that the substantial benefits of RSV vaccination outweigh these identified risks associated with vaccines.

**Key Points:** *Question:* Is there an increased risk of developing Guillain-Barré syndrome (GBS) following vaccination with RSVPreF3+AS01 (AREXVY®) and RSVPreF (ABRYSVO®) among Medicare beneficiaries aged 65 years and older?

*Findings:* In a self-controlled case series analysis of Medicare beneficiaries with medical record review, we estimated the incidence rate ratio (IRR) comparing GBS in pre-specified risk and control intervals following vaccination with RSVPreF3+AS01 (IRR: 2.46, 95%CI: 1.19-5.08) or RSVPreF (IRR: 2.02, 95%CI: 0.93-4.40). RSV vaccines were associated with fewer than 10 excess GBS cases per 1 million vaccine doses.

*Meaning:* These findings suggest an increased risk of GBS following RSV vaccination.

## 1. INTRODUCTION

Respiratory Syncytial Virus (RSV) infection can cause Lower Respiratory Tract Disease (LRTD), which may lead to life-threatening pneumonia and bronchiolitis.^1^ According to the Centers for Disease Control and Prevention (CDC), RSV infection causes approximately 60,000-160,000 hospitalizations and 6,000-10,000 deaths annually among adults 60 years of age and older.^1, 2^ The United States (U.S.) Food and Drug Administration (FDA) approved three RSV vaccines for prevention of LRTD caused by RSV infection in older adults.^3-5^ FDA approved the GlaxoSmithKline RSVPreF3+AS01 (AREXVY®) vaccine on May 03, 2023, the Pfizer RSVPreF (ABRYSVO®) vaccine on May 31, 2023, and the Moderna mRNA-1345 (mRESVIA®) on May 31, 2024 for use in adults aged 60 years and older.^3-5^ RSVPreF3+AS01 and RSVPreF were further approved for use in individuals 50-59 and 18-59 years of age, respectively, who are at increased risk for LRTD caused by RSV.^4, 6^ As of June 2024, the CDC’s Advisory Committee on Immunization Practices recommended that all adults aged 75 years and older and adults aged 60-74 years who are at an increased risk of severe RSV disease should receive a single dose of RSV vaccine.^2, 7^

The Center for Biologics Evaluation and Research within the FDA uses Centers for Medicare & Medicaid Services (CMS) administrative claims data to conduct active post-market surveillance to monitor the safety of vaccines and evaluate potential safety signals identified in pre-licensure clinical trials and passive surveillance systems.^8^ A small number of Guillain-Barré syndrome (GBS) cases in RSV-vaccinated and placebo groups were identified in safety data from clinical trials supporting licensure of RSVPreF3+AS01 and RSVPreF vaccines in adults aged 60 years and older.^9, 10^ Initial findings from reports submitted to the Vaccine Adverse Event Reporting System (VAERS) identified estimated GBS reporting rates after RSV vaccination (4.4 and 1.8 reports per million doses of RSVPreF and RSVPreF3+AS01 vaccines, respectively) higher than expected background rates in a vaccinated population.^11^ However, limitations in safety data accrued during these studies include generalizability, insufficient sample size to assess rare health outcomes, and reporting biases.^11^

Therefore, FDA conducted three separate post-market analyses to assess the risk of GBS following RSVPreF3+AS01 and RSVPreF vaccines among Medicare beneficiaries aged 65 years and older. A preliminary observed vs. expected (OvE) analysis compared the rates of GBS following either vaccine to historical control (expected) rates. Subsequently, more robust self-controlled case series (SCCS) analyses that adjusted for confounding were conducted to further evaluate the risk of GBS following the two RSV vaccines. An early-season SCCS analysis included beneficiaries vaccinated between May 2023 and October 2023, to support early detection of GBS risk shortly after the licensure of these vaccines. The end-of-season SCCS analysis included beneficiaries vaccinated between May 2023 and January 2024 and utilized medical record review (MRR) to provide more precise estimates of GBS risk following RSV vaccination.^12^ The findings presented in this manuscript focus on results from the end-of-season SCCS analysis.

## 2. METHODS

Post-market RSV vaccine monitoring utilized CMS administrative claims databases to assess the risk of GBS following RSV vaccination among Medicare beneficiaries enrolled in Medicare Fee-for-Service (FFS; Parts A and B) and Medicare Part D, using an SCCS study design.

### 2.1. Study Design, Data Sources, and Study Period

The SCCS design compares the incidence of GBS during periods of hypothesized excess risk due to vaccine administration (risk interval) to the incidence in the rest of the observation period (control interval). Only incident outcomes contribute to the SCCS analyses, and estimation of risk occurs within individuals adjusting for time-invariant confounding.^13^ The statistical analysis plan was pre-specified in a study protocol, and specifications are summarized in eTable 1.^12^

We used administrative claims data from Medicare FFS and Part D, accessed via the Shared Systems Data, capturing health services across inpatient, outpatient, and other community settings. The Medicare Enrollment Database was used to capture sociodemographic characteristics and death. Nursing home residency was captured from the Minimum Data Set 3.0.^14^ Socioeconomic deprivation of beneficiaries’ area of residence was assessed using the Area Deprivation Index.^15^

The study start date for the end-of-season SCCS analysis aligned with the earliest RSV vaccine approval date (May 3, 2023), and the study end date was July 13, 2024.^12^

### 2.2. Study Population, Exposure, Health Outcome, and Follow-up Times

The analysis included beneficiaries who were (i) aged 65 years and older enrolled in FFS and Part D on their first observed RSV vaccination date, and (ii) continuously enrolled in FFS 365 days prior to RSV vaccination.

Exposure was defined as receipt of a single dose of RSVPreF3+AS01 or RSVPreF vaccines, occurring after May 3, 2023 and May 31, 2023, respectively. Vaccinations were identified using Current Procedural Terminology/Healthcare Common Procedure Coding System codes or National Drug Codes. Persons with multiple RSV vaccine products administered on the same day, and persons who received RSV vaccination after the vaccination cutoff date were excluded from the study. The vaccination cutoff date was set as January 28, 2024 to ensure at least 90% data completeness at observation period end to ensure complete capture of GBS outcome.

GBS was identified using International Classification of Disease, 10^th^ Revision, Clinical Modification diagnosis code G61.0 in the primary diagnosis position on hospital inpatient claims. Days 1–42 post-vaccination were considered the risk interval while days 43-90 post-vaccination was the control interval. The selected risk interval was determined based on literature reviews and clinical consultation.^16^ GBS was required to be incident (i.e., no GBS outcome in the prior 365 days).

Persons were followed starting the day after vaccination and censored at the first of disenrollment, death, the end of the study period, or the end of the 90-day observation period. An example presenting observation period, risk, and control intervals for an individual with a qualifying GBS outcome following RSV vaccination is outlined in eFigure 1.^12^

### 2.3. Medical Record Review (MRR)

An MRR was conducted on available GBS cases that qualified for the SCCS analyses to validate the claims-based outcome definition. Each received record was independently adjudicated by two neurologists using the Brighton Collaboration’s case definition for GBS; cases classified as Brighton Levels 1, 2, or 3 of diagnostic certainty were classified as “chart-confirmed” GBS cases.^17^ Positive predictive values (PPVs) with corresponding 95% confidence intervals (CIs) were calculated as the percentage of received records classified as chart-confirmed. Results from this MRR were utilized to conduct inferential analyses.

### 2.4. Statistical Analyses

#### 2.4.1. Descriptive Analyses

We summarized characteristics of Medicare beneficiaries that received RSV vaccines by demographic, geographic, socioeconomic status, and concomitant vaccination status. Concomitant vaccination was defined as receipt of certain vaccines on the same date as RSV vaccination. Weekly vaccination uptake in the population was summarized.

#### 2.4.2. Inferential Analyses

We conducted the end-of-season SCCS analysis upon accumulation of a sufficient number of GBS cases to detect an incidence rate ratio (IRR) of at least 3 at 80% power with a two-sided alpha of 0.05.^12^ This analysis was conducted for (i) all claims-identified cases, (ii) chart-confirmed cases, and (iii) chart-confirmed cases and cases unreturned via MRR, with PPV-based imputation applied to unreturned cases, hereafter referred to as “PPV-based imputation analyses cases”. The PPV-based imputation adjustment utilized a quantitative bias analysis. Multiple datasets were created where the status of unreturned cases was imputed by assigning them the status of “chart-confirmed” with probability equal to the PPV. The primary analysis was repeated on multiple datasets and IRR estimates were pooled.^18^

For the primary analyses, a conditional Poisson regression was used to estimate IRRs and 95% CIs, comparing GBS rates in risk and control intervals for each RSV vaccine product. Attributable risk (AR) estimates and 95% CIs were calculated as the number of excess cases per 100,000 vaccine doses and per 100,000 person-years.^19^ Statistical significance was determined using two-sided hypothesis tests with an alpha of 0.05. Primary analyses included adjustments for outcome-dependent observation time (i.e., censoring of the observation period due to death or disenrollment; referred to as the Farrington adjustment), seasonality adjustment using incidence rates of GBS estimated from Medicare FFS population aged 65 years and older from corresponding calendar months in 2022-2023 used as baseline incidence of GBS in risk and control intervals, and PPV-based imputation adjustment to account for potential misclassification of GBS cases in administrative claims data.^18, 20^

Secondary analyses stratifying by concomitant vaccination status were conducted for each vaccine product with a statistically significant elevation of IRR in the primary analyses, to evaluate the potential effect of same-day concomitant vaccination.

We conducted sensitivity analyses that (i) incorporated a 14-day washout interval between risk and control intervals to evaluate potential bias associated with carryover effects from RSV vaccination contributing to the risk in control intervals, and (ii) included an individuals’ full observation period length for persons that died or disenrolled to evaluate potential bias from a violation of the SCCS assumption that observation length is independent of outcomes.^12, 13^

All analyses were conducted using R 4.3.4 (R Foundation for Statistical Computing, Vienna, Austria) and SAS v.9.4 (SAS Institute Inc. Cary, NC, United States).

This surveillance activity was conducted as part of FDA public health surveillance mandate. These safety surveillance activities are not considered research and therefore are exempt from institutional review board review and approval.^21^

## 3. RESULTS

### 3.1. Descriptive Results

We observed 3,226,689 Medicare beneficiaries, aged 65 years and older, that received RSV vaccination before January 28, 2024. Of these, 2,202,247 beneficiaries (68.25%) received RSVPreF3+AS01 vaccine and 1,024,442 beneficiaries (31.75%) received RSVPreF vaccine. Population characteristics were largely consistent by RSV vaccine products and are summarized in Table 1. Weekly vaccination uptake trends by RSV vaccine products are presented in Figure 1.

**Figure 1:**
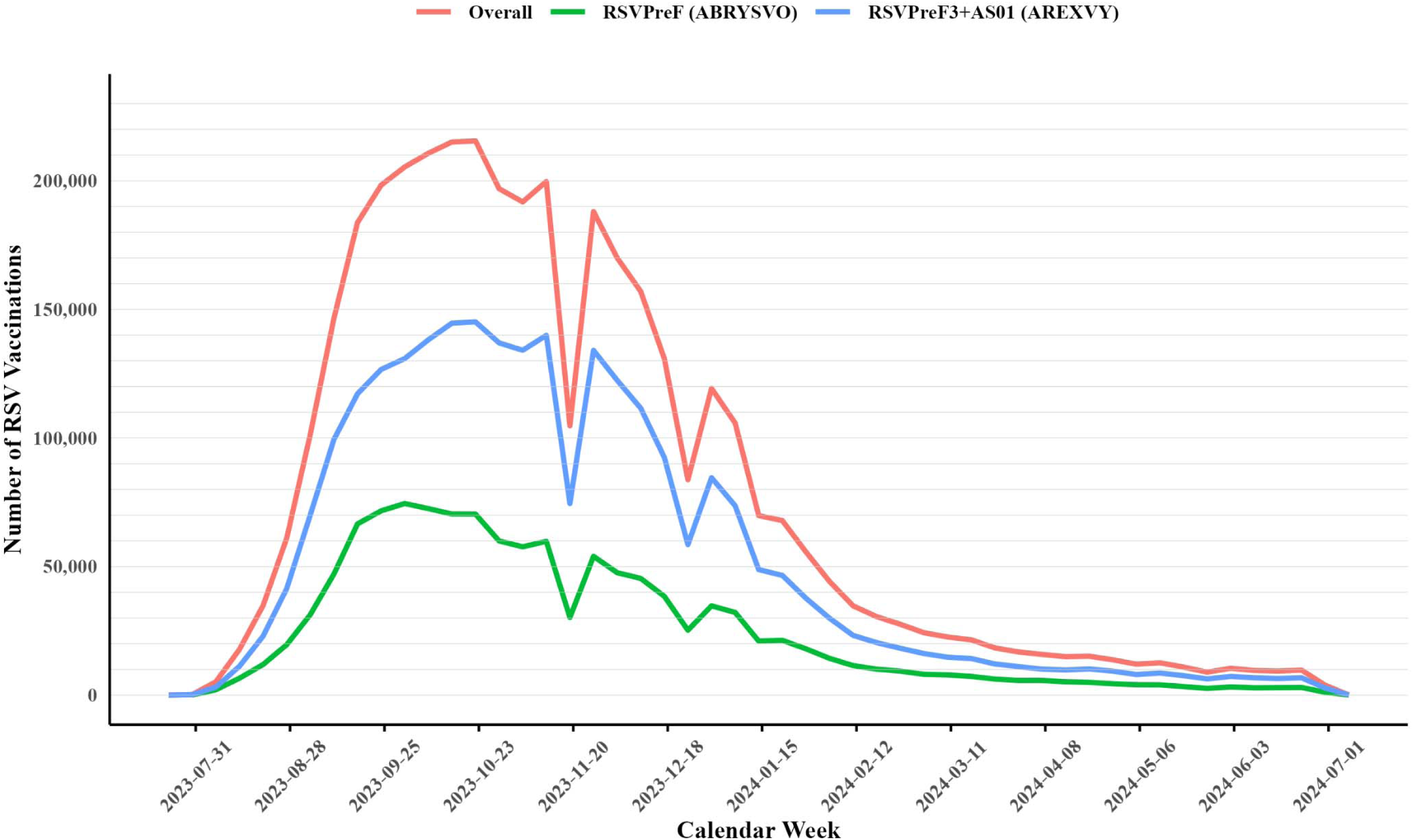
Weekly RSV Vaccination Uptake Trends Among Medicare Beneficiaries 65 Years and Older, Overall and by RSV Vaccine Product.

**Table 1:**
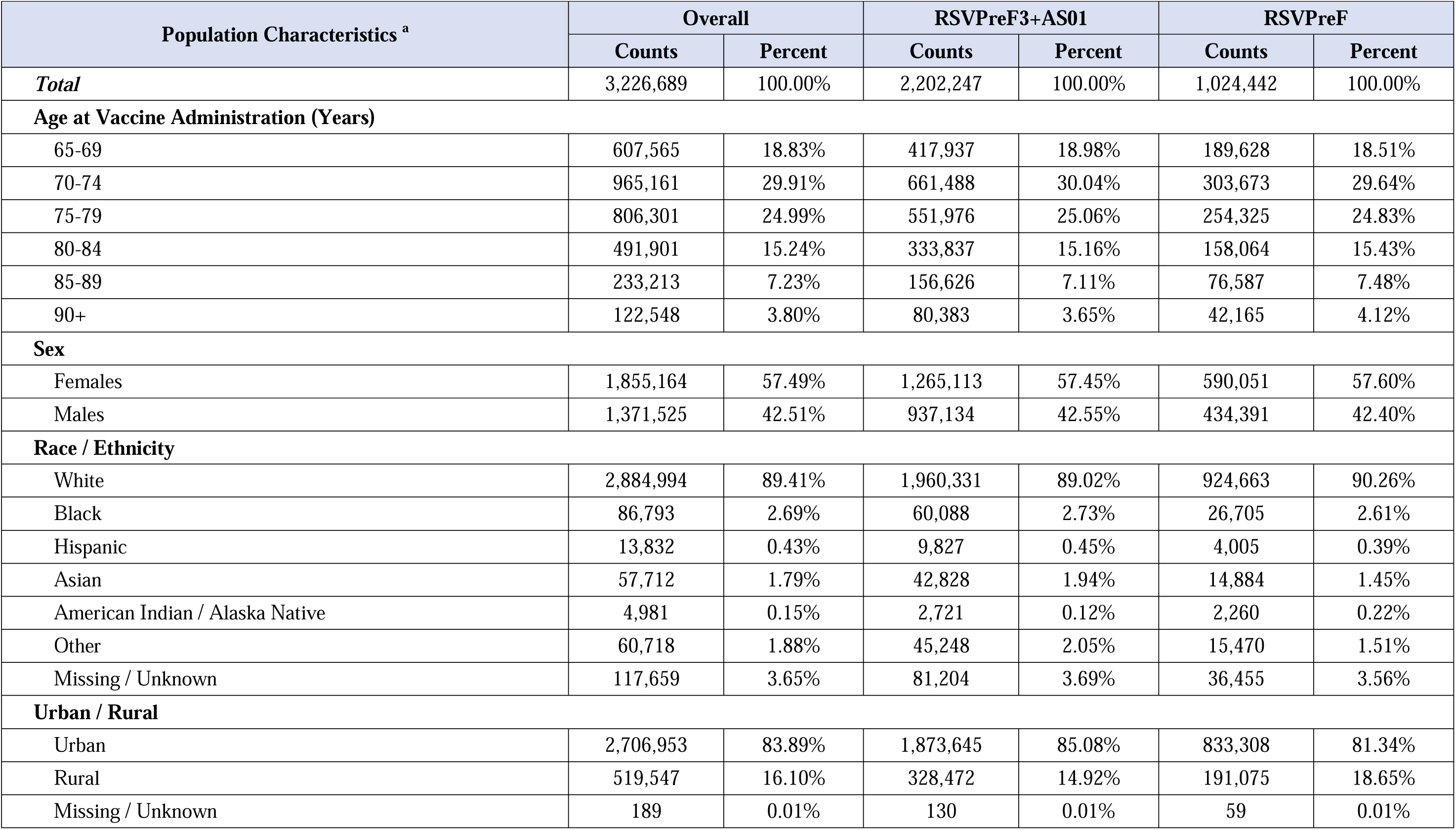
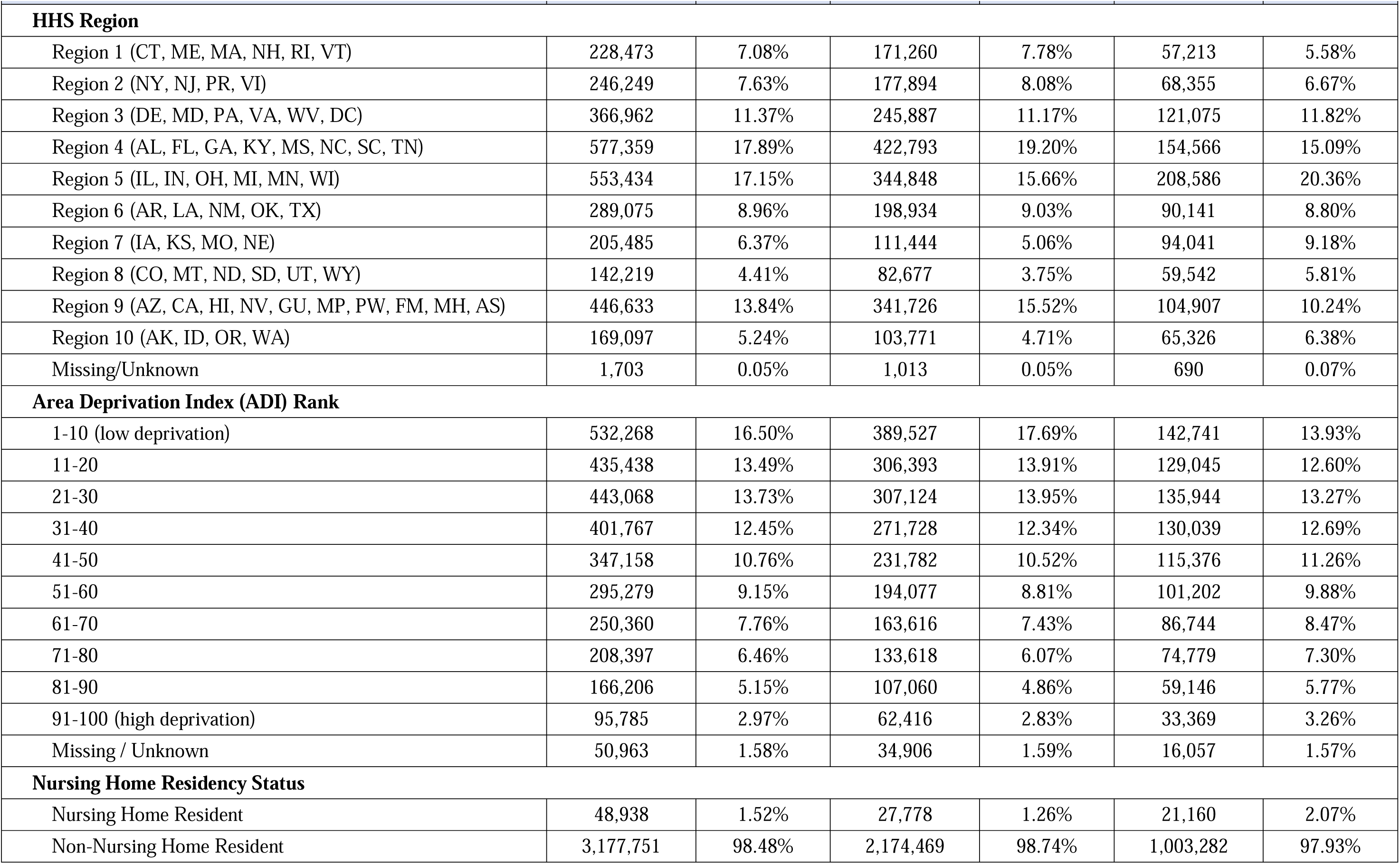
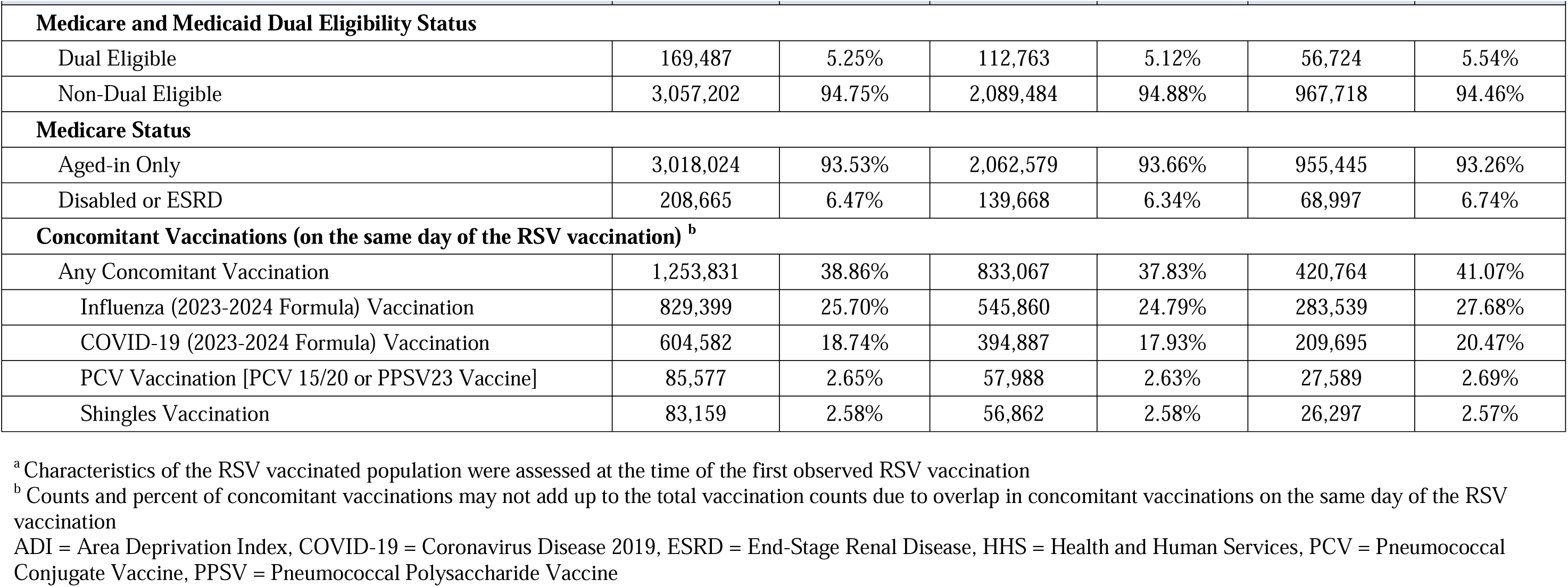
Characteristics of Medicare Beneficiaries 65 Years and Older that Received an RSV Vaccination and Eligible for GBS SCCS Analysis; Overall and By RSV Vaccine Product.

| Population Characteristics <sup>a</sup> | Overall |  | RSVPreF3+AS01 |  | RSVPreF |  |
| --- | --- | --- | --- | --- | --- | --- |
|  | Counts | Percent | Counts | Percent | Counts | Percent |
| **Total** | 3,226,689 | 100.00% | 2,202,247 | 100.00% | 1,024,442 | 100.00% |
| **Age at Vaccine Administration (Years)** |  |  |  |  |  |  |
| 65-69 | 607,565 | 18.83% | 417,937 | 18.98% | 189,628 | 18.51% |
| 70-74 | 965,161 | 29.91% | 661,488 | 30.04% | 303,673 | 29.64% |
| 75-79 | 806,301 | 24.99% | 551,976 | 25.06% | 254,325 | 24.83% |
| 80-84 | 491,901 | 15.24% | 333,837 | 15.16% | 158,064 | 15.43% |
| 85-89 | 233,213 | 7.23% | 156,626 | 7.11% | 76,587 | 7.48% |
| 90+ | 122,548 | 3.80% | 80,383 | 3.65% | 42,165 | 4.12% |
| **Sex** |  |  |  |  |  |  |
| Females | 1,855,164 | 57.49% | 1,265,113 | 57.45% | 590,051 | 57.60% |
| Males | 1,371,525 | 42.51% | 937,134 | 42.55% | 434,391 | 42.40% |
| **Race / Ethnicity** |  |  |  |  |  |  |
| White | 2,884,994 | 89.41% | 1,960,331 | 89.02% | 924,663 | 90.26% |
| Black | 86,793 | 2.69% | 60,088 | 2.73% | 26,705 | 2.61% |
| Hispanic | 13,832 | 0.43% | 9,827 | 0.45% | 4,005 | 0.39% |
| Asian | 57,712 | 1.79% | 42,828 | 1.94% | 14,884 | 1.45% |
| American Indian / Alaska Native | 4,981 | 0.15% | 2,721 | 0.12% | 2,260 | 0.22% |
| Other | 60,718 | 1.88% | 45,248 | 2.05% | 15,470 | 1.51% |
| Missing / Unknown | 117,659 | 3.65% | 81,204 | 3.69% | 36,455 | 3.56% |
| **Urban / Rural** |  |  |  |  |  |  |
| Urban | 2,706,953 | 83.89% | 1,873,645 | 85.08% | 833,308 | 81.34% |
| Rural | 519,547 | 16.10% | 328,472 | 14.92% | 191,075 | 18.65% |
| Missing / Unknown | 189 | 0.01% | 130 | 0.01% | 59 | 0.01% |
|  | Counts | Percent | Counts | Percent | Counts | Percent |
| **HHS Region** |  |  |  |  |  |  |
| Region 1 (CT, ME, MA, NH, RI, VT) | 228,473 | 7.08% | 171,260 | 7.78% | 57,213 | 5.58% |
| Region 2 (NY, NJ, PR, VI) | 246,249 | 7.63% | 177,894 | 8.08% | 68,355 | 6.67% |
| Region 3 (DE, MD, PA, VA, WV, DC) | 366,962 | 11.37% | 245,887 | 11.17% | 121,075 | 11.82% |
| Region 4 (AL, FL, GA, KY, MS, NC, SC, TN) | 577,359 | 17.89% | 422,793 | 19.20% | 154,566 | 15.09% |
| Region 5 (IL, IN, OH, MI, MN, WI) | 553,434 | 17.15% | 344,848 | 15.66% | 208,586 | 20.36% |
| Region 6 (AR, LA, NM, OK, TX) | 289,075 | 8.96% | 198,934 | 9.03% | 90,141 | 8.80% |
| Region 7 (IA, KS, MO, NE) | 205,485 | 6.37% | 111,444 | 5.06% | 94,041 | 9.18% |
| Region 8 (CO, MT, ND, SD, UT, WY) | 142,219 | 4.41% | 82,677 | 3.75% | 59,542 | 5.81% |
| Region 9 (AZ, CA, HI, NV, GU, MP, PW, FM, MH, AS) | 446,633 | 13.84% | 341,726 | 15.52% | 104,907 | 10.24% |
| Region 10 (AK, ID, OR, WA) | 169,097 | 5.24% | 103,771 | 4.71% | 65,326 | 6.38% |
| Missing/Unknown | 1,703 | 0.05% | 1,013 | 0.05% | 690 | 0.07% |
| **Area Deprivation Index (ADI) Rank** |  |  |  |  |  |  |
| 1-10 (low deprivation) | 532,268 | 16.50% | 389,527 | 17.69% | 142,741 | 13.93% |
| 11-20 | 435,438 | 13.49% | 306,393 | 13.91% | 129,045 | 12.60% |
| 21-30 | 443,068 | 13.73% | 307,124 | 13.95% | 135,944 | 13.27% |
| 31-40 | 401,767 | 12.45% | 271,728 | 12.34% | 130,039 | 12.69% |
| 41-50 | 347,158 | 10.76% | 231,782 | 10.52% | 115,376 | 11.26% |
| 51-60 | 295,279 | 9.15% | 194,077 | 8.81% | 101,202 | 9.88% |
| 61-70 | 250,360 | 7.76% | 163,616 | 7.43% | 86,744 | 8.47% |
| 71-80 | 208,397 | 6.46% | 133,618 | 6.07% | 74,779 | 7.30% |
| 81-90 | 166,206 | 5.15% | 107,060 | 4.86% | 59,146 | 5.77% |
| 91-100 (high deprivation) | 95,785 | 2.97% | 62,416 | 2.83% | 33,369 | 3.26% |
| Missing / Unknown | 50,963 | 1.58% | 34,906 | 1.59% | 16,057 | 1.57% |
| **Nursing Home Residency Status** |  |  |  |  |  |  |
| Nursing Home Resident | 48,938 | 1.52% | 27,778 | 1.26% | 21,160 | 2.07% |
| Non-Nursing Home Resident | 3,177,751 | 98.48% | 2,174,469 | 98.74% | 1,003,282 | 97.93% |
|  | Counts | Percent | Counts | Percent | Counts | Percent |
| **Medicare and Medicaid Dual Eligibility Status** |  |  |  |  |  |  |
| Dual Eligible | 169,487 | 5.25% | 112,763 | 5.12% | 56,724 | 5.54% |
| Non-Dual Eligible | 3,057,202 | 94.75% | 2,089,484 | 94.88% | 967,718 | 94.46% |
| **Medicare Status** |  |  |  |  |  |  |
| Aged-in Only | 3,018,024 | 93.53% | 2,062,579 | 93.66% | 955,445 | 93.26% |
| Disabled or ESRD | 208,665 | 6.47% | 139,668 | 6.34% | 68,997 | 6.74% |
| **Concomitant Vaccinations (on the same day of the RSV vaccination) <sup>b</sup>** |  |  |  |  |  |  |
| Any Concomitant Vaccination | 1,253,831 | 38.86% | 833,067 | 37.83% | 420,764 | 41.07% |
| Influenza (2023-2024 Formula) Vaccination | 829,399 | 25.70% | 545,860 | 24.79% | 283,539 | 27.68% |
| COVID-19 (2023-2024 Formula) Vaccination | 604,582 | 18.74% | 394,887 | 17.93% | 209,695 | 20.47% |
| PCV Vaccination [PCV 15/20 or PPSV23 Vaccine] | 85,577 | 2.65% | 57,988 | 2.63% | 27,589 | 2.69% |
| Shingles Vaccination | 83,159 | 2.58% | 56,862 | 2.58% | 26,297 | 2.57% |

<sup>a</sup> Characteristics of the RSV vaccinated population were assessed at the time of the first observed RSV vaccination
<sup>b</sup> Counts and percent of concomitant vaccinations may not add up to the total vaccination counts due to overlap in concomitant vaccinations on the same day of the RSV vaccination
ADI = Area Deprivation Index, COVID-19 = Coronavirus Disease 2019, ESRD = End-Stage Renal Disease, HHS = Health and Human Services, PCV = Pneumococcal Conjugate Vaccine, PPSV = Pneumococcal Polysaccharide Vaccine

### 3.2. Medical Record Review Results

We requested records for all 95 incident GBS cases identified following RSV vaccination using the claims-based definition; records from 75 cases were received and adjudicated. Table 2 summarizes the medical records status, case classifications derived after adjudication of records, and PPVs of GBS following RSV vaccination. The overall PPV of the claims-based GBS definition in the primary diagnosis position on hospital inpatient claims following RSV vaccination was slightly lower (PPV: 68.00% [95% CI: 56.79, 77.46]) compared to a prior MRR of GBS following influenza vaccination (PPV: 71.21% [95% CI: 63.49, 78.94]).^22^ PPVs of GBS following RSV vaccination were observed to be higher for the control interval (PPV: 81.82% [95% CI: 61.48, 92.69]) compared to the risk interval (PPV: 62.26% [95% CI: 48.81, 74.06]) (Table 2).

**Table 2:**
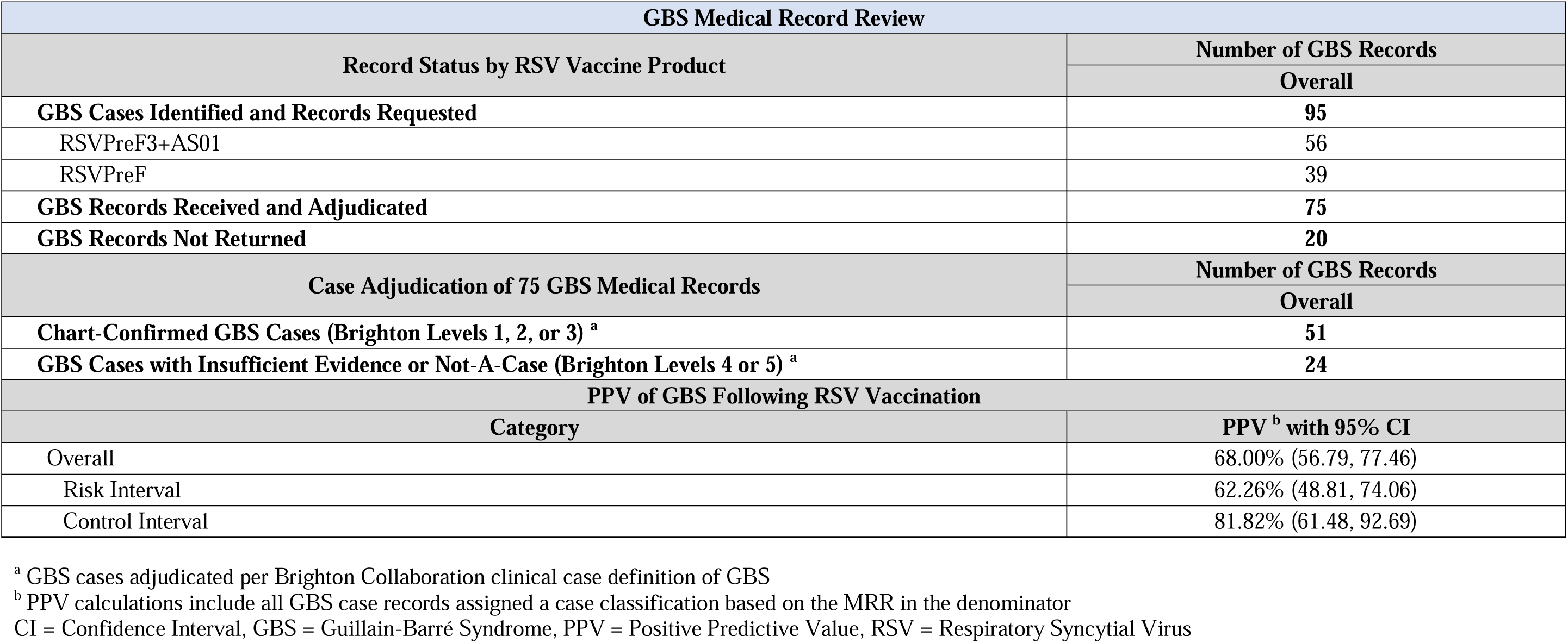
Summary of GBS Medical Record Review (MRR) Results and Positive Predictive Value (PPV) with 95% Confidence Intervals.

| GBS Medical Record Review |  |
| --- | --- |
| Record Status by RSV Vaccine Product | Number of GBS Records |
|  | Overall |
| **GBS Cases Identified and Records Requested** | **95** |
| RSVPreF3+AS01 | 56 |
| RSVPreF | 39 |
| **GBS Records Received and Adjudicated** | **75** |
| **GBS Records Not Returned** | **20** |
| Case Adjudication of 75 GBS Medical Records | Number of GBS Records |
|  | Overall |
| **Chart-Confirmed GBS Cases (Brighton Levels 1, 2, or 3) <sup>a</sup>** | **51** |
| **GBS Cases with Insufficient Evidence or Not-A-Case (Brighton Levels 4 or 5) <sup>a</sup>** | **24** |
| PPV of GBS Following RSV Vaccination |  |
| Category | PPV <sup>b</sup> with 95% CI |
| Overall | 68.00% (56.79, 77.46) |
| Risk Interval | 62.26% (48.81, 74.06) |
| Control Interval | 81.82% (61.48, 92.69) |

<sup>a</sup> GBS cases adjudicated per Brighton Collaboration clinical case definition of GBS
<sup>b</sup> PPV calculations include all GBS case records assigned a case classification based on the MRR in the denominator
CI = Confidence Interval, GBS = Guillain-Barré Syndrome, PPV = Positive Predictive Value, RSV = Respiratory Syncytial Virus

### 3.3. Inferential Results

Results from primary, secondary, and sensitivity SCCS analyses of GBS following RSV vaccination are summarized below.

For the primary analyses, a total of 95 incident cases were identified: 56 following RSVPreF3+AS01 and 39 following RSVPreF vaccination. Table 3 summarizes the IRR and AR estimates from primary analyses of GBS following RSV vaccination for (i) PPV-based imputation analyses cases (n=71), (ii) chart-confirmed cases (n=51), and (iii) all claims-identified cases (n=95).

**Table 3:**
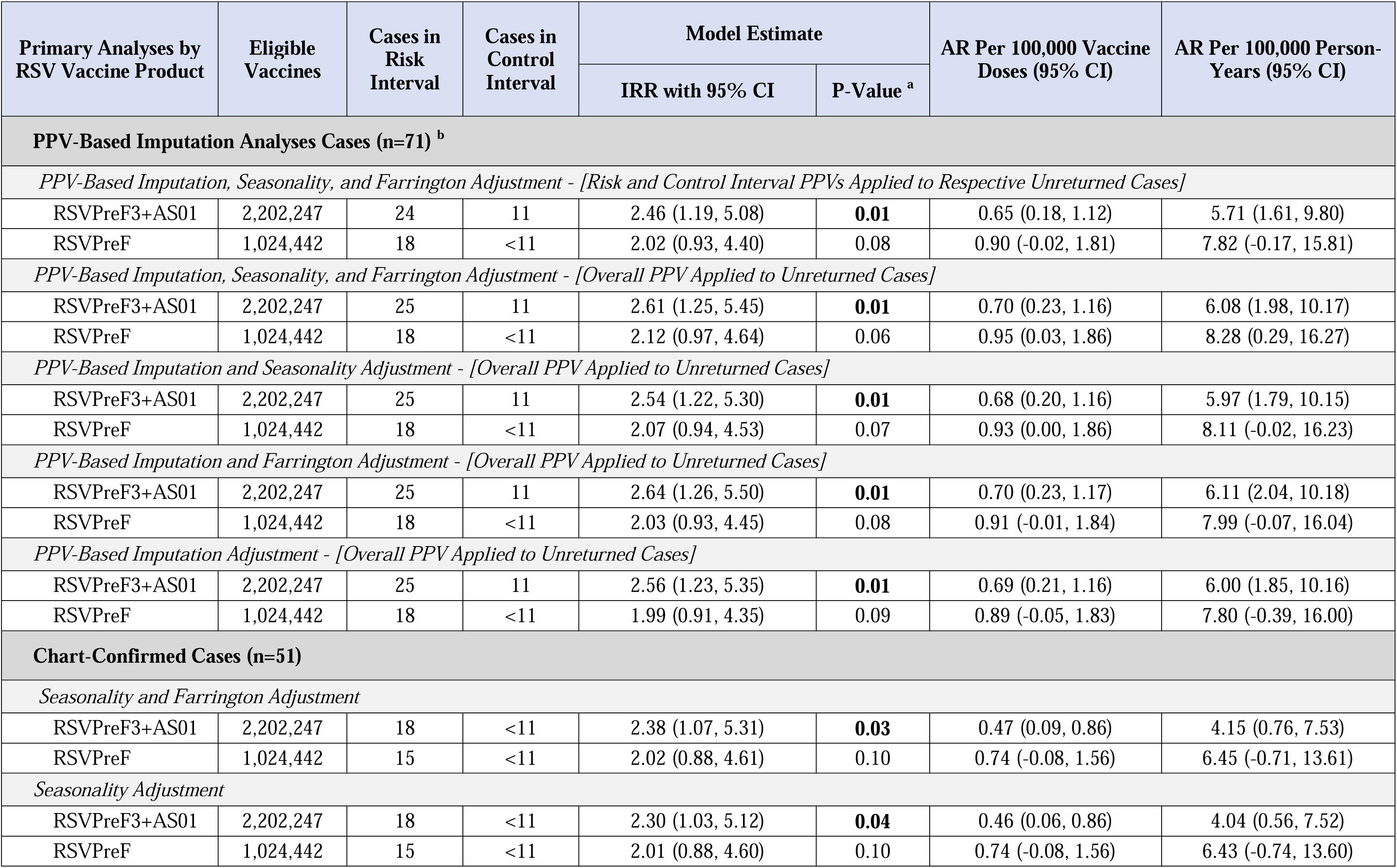
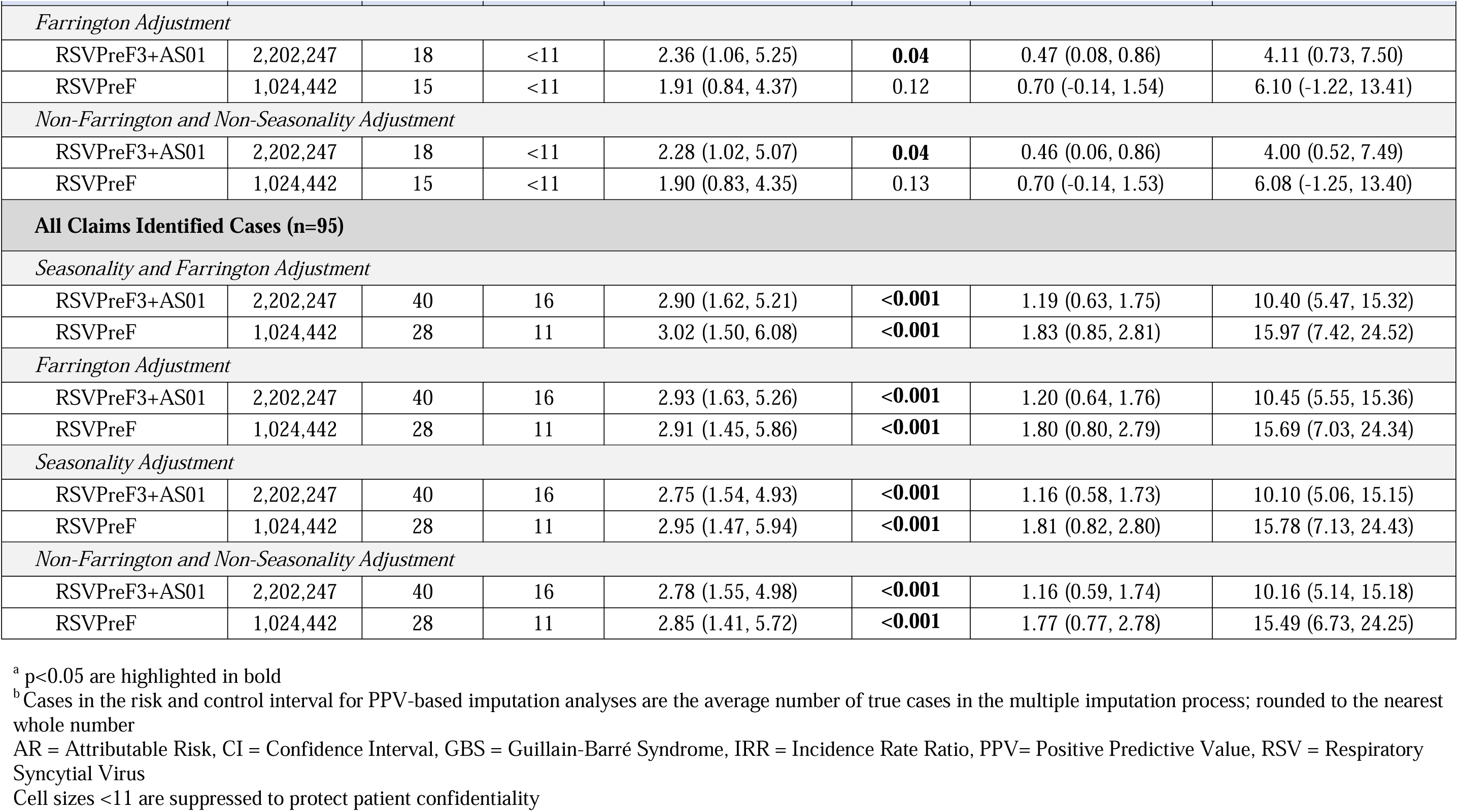
Primary Analyses: IRR and AR with Corresponding 95% CIs of GBS following RSV Vaccination for PPV-Based Imputation Analyses, Chart-Confirmed, and All Claims-Identified GBS Cases.

| Primary Analyses by RSV Vaccine Product | Eligible Vaccines | Cases in Risk Interval | Cases in Control Interval | Model Estimate |  | AR Per 100,000 Vaccine Doses (95% CI) | AR Per 100,000 Person-Years (95% CI) |
| --- | --- | --- | --- | --- | --- | --- | --- |
|  |  |  |  | IRR with 95% CI | P-Value <sup>a</sup> |  |  |
| PPV-Based Imputation Analyses Cases (n=71) <sup>b</sup> |  |  |  |  |  |  |  |
| PPV-Based Imputation, Seasonality, and Farrington Adjustment - [Risk and Control Interval PPVs Applied to Respective Unreturned Cases] |  |  |  |  |  |  |  |
| RSVPreF3+AS01 | 2,202,247 | 24 | 11 | 2.46 (1.19, 5.08) | 0.01 | 0.65 (0.18, 1.12) | 5.71 (1.61, 9.80) |
| RSVPreF | 1,024,442 | 18 | <11 | 2.02 (0.93, 4.40) | 0.08 | 0.90 (-0.02, 1.81) | 7.82 (-0.17, 15.81) |
| PPV-Based Imputation, Seasonality, and Farrington Adjustment - [Overall PPV Applied to Unreturned Cases] |  |  |  |  |  |  |  |
| RSVPreF3+AS01 | 2,202,247 | 25 | 11 | 2.61 (1.25, 5.45) | 0.01 | 0.70 (0.23, 1.16) | 6.08 (1.98, 10.17) |
| RSVPreF | 1,024,442 | 18 | <11 | 2.12 (0.97, 4.64) | 0.06 | 0.95 (0.03, 1.86) | 8.28 (0.29, 16.27) |
| PPV-Based Imputation and Seasonality Adjustment - [Overall PPV Applied to Unreturned Cases] |  |  |  |  |  |  |  |
| RSVPreF3+AS01 | 2,202,247 | 25 | 11 | 2.54 (1.22, 5.30) | 0.01 | 0.68 (0.20, 1.16) | 5.97 (1.79, 10.15) |
| RSVPreF | 1,024,442 | 18 | <11 | 2.07 (0.94, 4.53) | 0.07 | 0.93 (0.00, 1.86) | 8.11 (-0.02, 16.23) |
| PPV-Based Imputation and Farrington Adjustment - [Overall PPV Applied to Unreturned Cases] |  |  |  |  |  |  |  |
| RSVPreF3+AS01 | 2,202,247 | 25 | 11 | 2.64 (1.26, 5.50) | 0.01 | 0.70 (0.23, 1.17) | 6.11 (2.04, 10.18) |
| RSVPreF | 1,024,442 | 18 | <11 | 2.03 (0.93, 4.45) | 0.08 | 0.91 (-0.01, 1.84) | 7.99 (-0.07, 16.04) |
| PPV-Based Imputation Adjustment - [Overall PPV Applied to Unreturned Cases] |  |  |  |  |  |  |  |
| RSVPreF3+AS01 | 2,202,247 | 25 | 11 | 2.56 (1.23, 5.35) | 0.01 | 0.69 (0.21, 1.16) | 6.00 (1.85, 10.16) |
| RSVPreF | 1,024,442 | 18 | <11 | 1.99 (0.91, 4.35) | 0.09 | 0.89 (-0.05, 1.83) | 7.80 (-0.39, 16.00) |
| Chart-Confirmed Cases (n=51) |  |  |  |  |  |  |  |
| Seasonality and Farrington Adjustment |  |  |  |  |  |  |  |
| RSVPreF3+AS01 | 2,202,247 | 18 | <11 | 2.38 (1.07, 5.31) | 0.03 | 0.47 (0.09, 0.86) | 4.15 (0.76, 7.53) |
| RSVPreF | 1,024,442 | 15 | <11 | 2.02 (0.88, 4.61) | 0.10 | 0.74 (-0.08, 1.56) | 6.45 (-0.71, 13.61) |
| Seasonality Adjustment |  |  |  |  |  |  |  |
| RSVPreF3+AS01 | 2,202,247 | 18 | <11 | 2.30 (1.03, 5.12) | 0.04 | 0.46 (0.06, 0.86) | 4.04 (0.56, 7.52) |
| RSVPreF | 1,024,442 | 15 | <11 | 2.01 (0.88, 4.60) | 0.10 | 0.74 (-0.08, 1.56) | 6.43 (-0.74, 13.60) |
|  |  |  |  | IRR with 95% CI | P-Value <sup>a</sup> |  |  |
| Farrington Adjustment |  |  |  |  |  |  |  |
| RSVPreF3+AS01 | 2,202,247 | 18 | <11 | 2.36 (1.06, 5.25) | 0.04 | 0.47 (0.08, 0.86) | 4.11 (0.73, 7.50) |
| RSVPreF | 1,024,442 | 15 | <11 | 1.91 (0.84, 4.37) | 0.12 | 0.70 (-0.14, 1.54) | 6.10 (-1.22, 13.41) |
| Non-Farrington and Non-Seasonality Adjustment |  |  |  |  |  |  |  |
| RSVPreF3+AS01 | 2,202,247 | 18 | <11 | 2.28 (1.02, 5.07) | 0.04 | 0.46 (0.06, 0.86) | 4.00 (0.52, 7.49) |
| RSVPreF | 1,024,442 | 15 | <11 | 1.90 (0.83, 4.35) | 0.13 | 0.70 (-0.14, 1.53) | 6.08 (-1.25, 13.40) |
| All Claims Identified Cases (n=95) |  |  |  |  |  |  |  |
| Seasonality and Farrington Adjustment |  |  |  |  |  |  |  |
| RSVPreF3+AS01 | 2,202,247 | 40 | 16 | 2.90 (1.62, 5.21) | <0.001 | 1.19 (0.63, 1.75) | 10.40 (5.47, 15.32) |
| RSVPreF | 1,024,442 | 28 | 11 | 3.02 (1.50, 6.08) | <0.001 | 1.83 (0.85, 2.81) | 15.97 (7.42, 24.52) |
| Farrington Adjustment |  |  |  |  |  |  |  |
| RSVPreF3+AS01 | 2,202,247 | 40 | 16 | 2.93 (1.63, 5.26) | <0.001 | 1.20 (0.64, 1.76) | 10.45 (5.55, 15.36) |
| RSVPreF | 1,024,442 | 28 | 11 | 2.91 (1.45, 5.86) | <0.001 | 1.80 (0.80, 2.79) | 15.69 (7.03, 24.34) |
| Seasonality Adjustment |  |  |  |  |  |  |  |
| RSVPreF3+AS01 | 2,202,247 | 40 | 16 | 2.75 (1.54, 4.93) | <0.001 | 1.16 (0.58, 1.73) | 10.10 (5.06, 15.15) |
| RSVPreF | 1,024,442 | 28 | 11 | 2.95 (1.47, 5.94) | <0.001 | 1.81 (0.82, 2.80) | 15.78 (7.13, 24.43) |
| Non-Farrington and Non-Seasonality Adjustment |  |  |  |  |  |  |  |
| RSVPreF3+AS01 | 2,202,247 | 40 | 16 | 2.78 (1.55, 4.98) | <0.001 | 1.16 (0.59, 1.74) | 10.16 (5.14, 15.18) |
| RSVPreF | 1,024,442 | 28 | 11 | 2.85 (1.41, 5.72) | <0.001 | 1.77 (0.77, 2.78) | 15.49 (6.73, 24.25) |

<sup>a</sup> p<0.05 are highlighted in bold
<sup>b</sup> Cases in the risk and control interval for PPV-based imputation analyses are the average number of true cases in the multiple imputation process; rounded to the nearest whole number
AR = Attributable Risk, CI = Confidence Interval, GBS = Guillain-Barré Syndrome, IRR = Incidence Rate Ratio, PPV= Positive Predictive Value, RSV = Respiratory Syncytial Virus
Cell sizes <11 are suppressed to protect patient confidentiality

For PPV-based imputation analyses cases, a statistically significant elevation in risk was observed following RSVPreF3+AS01 vaccination (IRR: 2.46, [95% CI: 1.19, 5.08]) in seasonality, Farrington, and PPV-based imputation adjustment analysis that utilized separate risk and control interval PPVs on unreturned cases (Table 3). This elevation in GBS risk following RSVPreF3+AS01 remained statistically significant for: (i) PPV-based imputation analyses cases that utilized an overall PPV on unreturned cases, (ii) chart-confirmed cases, and (iii) all claims-identified cases (Table 3).

Although a statistically significant elevation in risk was observed following RSVPreF for all claims-identified cases, subsequent analyses showed an elevated yet non-statistically significant result (Table 3). A non-statistically significant elevation in risk was observed following RSVPreF vaccination (IRR: 2.02, [95% CI: 0.93, 4.40]) for PPV-based imputation analyses cases in seasonality, Farrington, and PPV-based imputation adjustment analysis that utilized separate risk and control interval PPVs on unreturned cases (Table 3). This result remained consistent for: (i) PPV-based imputation analyses cases that utilized an overall PPV on unreturned cases, and (ii) chart-confirmed cases (Table 3).

The AR per 100,000 doses following RSVPreF3+AS01 and RSVPreF were similar in magnitude (AR: 0.65, [95% CI: 0.18, 1.12] and AR: 0.90, [95% CI: -0.02, 1.81], respectively) in the most-adjusted analysis (i.e., seasonality, Farrington, and PPV-based imputation adjustment) that utilized separate risk and control interval PPVs on unreturned cases (Table 3). Figure 2 summarizes primary SCCS analyses results of GBS for: (i) PPV-based imputation analyses cases, (ii) chart-confirmed cases, and (iii) all claims-identified cases, following RSV vaccination.

**Figure 2:**
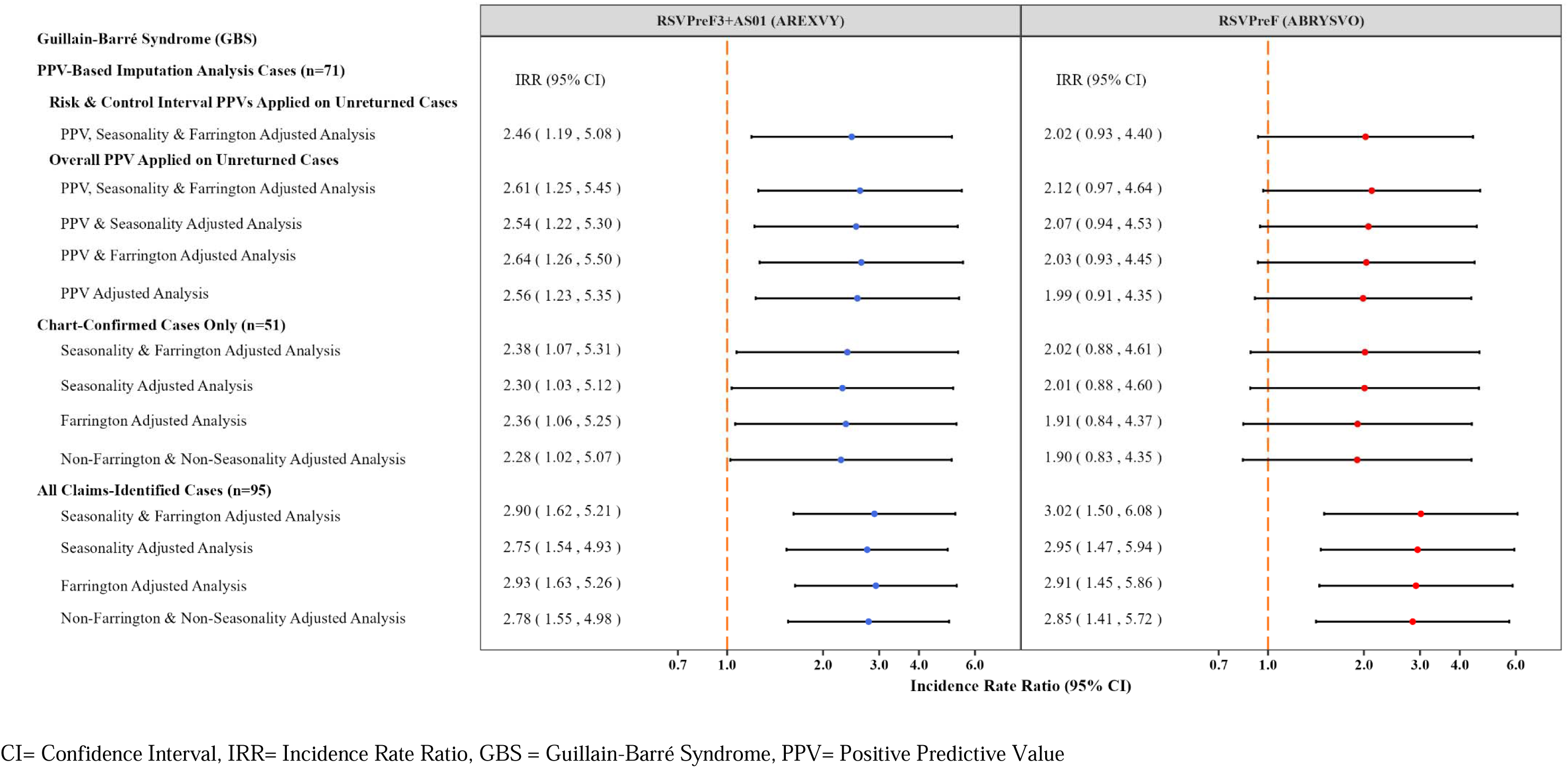
Primary Analyses: IRR with Corresponding 95% CIs of GBS following RSV Vaccination for PPV-Based Imputation Analyses, Chart-Confirmed, and All Claims-Identified GBS Cases.

In the secondary analyses, 20 (35.71%) and 19 (48.71%) GBS cases were observed to have a concomitant vaccination with RSVPreF3+AS01 and RSVPreF vaccines, respectively. eTable 2 and eFigure 3 summarize the IRR and AR estimates of GBS following RSV vaccination by concomitant vaccination status for all claims-identified cases. A statistically significant elevation in risk was observed following RSVPreF3+AS01 (IRR: 3.47, [95% CI: 1.61, 7.46]) and RSVPreF (IRR: 4.48, [95% CI: 1.50, 13.42]) without concomitant vaccination, whereas a non-statistically significant elevation in risk was observed following both RSV vaccines with concomitant vaccination in seasonality and Farrington-adjusted analyses {RSVPreF3+AS01: (IRR: 2.19, [95% CI: 0.87, 5.49]); RSVPreF: (IRR: 2.26, [95% CI: 0.89, 5.73])} (eTable 2). eTable 3 and eFigure 4 summarize the IRR and AR estimates of GBS following RSV vaccination by concomitant vaccination status for PPV-based imputation analyses cases, and chart-confirmed cases.

Sensitivity analyses results are presented in eTable 4 and eFigure 5. In an assessment of the washout interval of PPV-based imputation analyses cases, a non-statistically significant elevation in risk was observed following both RSV vaccines in the most-adjusted analysis {RSVPreF3+AS01: (IRR: 2.03, [95% CI: 0.93, 4.42]); RSVPreF: (IRR: 1.54, [95% CI: 0.69, 3.44])} (eTable 3). In the full-planned observation time analysis of PPV-based imputation analyses cases, a statistically significant elevation in risk was observed following RSVPreF3+AS01 (IRR: 2.40, [95% CI: 1.16, 4.95]) and risk was elevated yet not statistically significant following RSVPreF (IRR: 2.02, [95% CI: 0.93, 4.39]) in the most adjusted analysis (eTable 4).

Results from the early-season SCCS analysis are summarized in eTable 5 and eFigure 6, and results from the OvE analysis are summarized in eTable 6.

## 4. DISCUSSION

In our analysis of approximately 3.23 million RSV vaccinations among Medicare beneficiaries aged 65 years and older, we identified an elevated risk of GBS following the RSVPreF3+AS01 and RSVPreF vaccination. The risk of GBS following RSVPreF3+AS01 vaccination was elevated and statistically significant in the primary analyses and remained statistically significant with additional adjustments. Following RSVPreF vaccination, GBS risk was elevated although not statistically significant; these results remained consistent with additional adjustments. Our AR estimate indicates that approximately 6.5 excess GBS cases per 1 million doses were associated with RSVPreF3+AS01 and 9 GBS cases per 1 million doses were associated with RSVPreF. In secondary analyses of concomitant vaccinations, there was no evidence of difference in GBS risk among persons with and without concomitant vaccination with RSV vaccines. Although the uncertainty is high in this subgroup analysis due to low case counts, this may suggest that the elevation in GBS risk overall was not due to other vaccines administered on the same day.

These results are consistent with findings from other safety studies of the RSV vaccines as well as literature on the risk of GBS after other vaccines. Pre-licensure clinical trials and initial case reports of RSV vaccines have reported a small number of GBS cases following receipt of RSV vaccines.^9, 10^ Initial findings from reports submitted to VAERS, a U.S. passive surveillance system, were consistent with patterns observed in clinical trials, with GBS reporting rates after RSV vaccination (4.4 and 1.8 reports per million doses of RSVPreF and RSVPreF3+AS01 vaccines, respectively) being higher than expected background rates in a vaccinated population.^11^ Other vaccines, such as the influenza and recombinant zoster vaccines, have also been associated with an increased GBS risk, with less than one excess GBS case per million influenza vaccines in 8–21-day and 1–42-day post-vaccination, and 3 cases per million zoster vaccines in 1–42 days post-vaccination.^23, 24^ Although the excess cases of GBS post-RSV vaccination appears higher compared to these other vaccines, this risk is rare (i.e., fewer than 10 cases per million RSV vaccinations).

There are several strengths of this study. First, the SCCS design inherently adjusts for potential time invariant confounders such as sex and race/ethnicity, because the comparison occurs within, rather than between, individuals. Second, the study used data for all Medicare FFS beneficiaries, almost 30 million beneficiaries to produce reliable and generalizable estimates of GBS risk in this population. Third, access to medical records offered the ability to evaluate GBS cases identified in the claims database and adjust for potential misclassification of these cases.^13^ We used MRR to confirm GBS cases identified via claims-based definition per Brighton Classification case definition, and estimated PPV by risk and control intervals. By conducting analyses that restricted to chart-confirmed cases, and additionally incorporating interval-specific PPVs on unreturned cases, we improved the accuracy of our GBS risk estimates.

Despite these strengths, our study has several limitations. First, potential misspecification of post-RSV vaccination risk and control intervals for GBS could bias the estimates in either direction. Second, the estimates of GBS following RSV vaccination for the chart-confirmed and PPV-based imputation analyses cases in various analyses may be sensitive to the number of medical records received and adjudicated via MRR. Third, the SCCS design compares risk of a given outcome within individuals and is thus not meant to compare the risk of a given outcome between the vaccine products. Fourth, our study had limited power to conduct subgroup analyses by certain sociodemographic or geographic characteristics due to low GBS case counts within those stratifications. Finally, because residual and unmeasured confounding in observational studies cannot be ruled out, these results carry a level of uncertainty.

## 5. CONCLUSION

Our findings suggest an increased GBS risk following RSVPreF3+AS01 and RSVPreF among adults aged 65 years and older, with fewer than 10 excess cases per 1 million RSV vaccine doses. FDA continues to receive and adjudicate additional GBS medical records, and conduct additional analyses upon data accrual to provide more robust estimates. FDA maintains that these results support the safety profile of RSV vaccines administered to U.S. adults aged 65 years and older and that the substantial benefits of RSV vaccination in preventing RSV hospitalizations and LRTD outweigh potential risks associated with vaccines.

## Data Availability

Data from this study will not be shared.

## Abbreviations

AR: Attributable Risk
CDC: Centers for Disease Control and Prevention
CI: Confidence Interval
CMS: Centers for Medicare & Medicaid Services
FDA: Food and Drug Administration
FFS: Fee-for-Service
GBS: Guillain-Barré syndrome
IRR: Incidence Rate Ratio
LRTD: Lower Respiratory Tract Disease
MRR: Medical Record Review
OvE: Observed vs. Expected
PPV: Positive Predictive Value
RSV: Respiratory Syncytial Virus
SCCS: Self-Controlled Case Series
US: United States
VAERS: Vaccine Adverse Event Reporting System

## 6. ARTICLE INFORMATION

### Corresponding Author

Patricia C. Lloyd, PhD, ScM, Health Statistician, Office of Biostatistics and Pharmacovigilance, Center for Biologics Evaluation and Research, U.S. Food & Drug Administration, 10903 New Hampshire Ave., Building 71, Silver Spring, MD 20993

### Author Contributions

*Concept and design: Patricia C. Lloyd, Purva B. Shah, Henry T. Zhang, Nimesh Shah, Zhiruo Wan, Mao Hu, Meng Chen, Jing Wang, Yue Wu, Yoganand Chillarige, Richard A. Forshee, Steven A. Anderson*

*Acquisition, analysis, or interpretation of data: Patricia C. Lloyd, Purva B. Shah, Henry T. Zhang, Nimesh Shah, Narayan Nair, Zhiruo Wan, Mao Hu, Tainya C. Clarke, Meng Chen, Xinxin Lin, Rose Do, Jing Wang, Yue Wu, Yoganand Chillarige, Richard A. Forshee, Steven A. Anderson*

*Drafting of the manuscript: Patricia C. Lloyd, Purva B. Shah, Henry T. Zhang, Mao Hu, Yoganand Chillarige*

*Critical revision of the manuscript for important intellectual content: Patricia C. Lloyd, Purva B. Shah, Henry T. Zhang, Mao Hu, Yoganand Chillarige, Richard A. Forshee, Steven A. Anderson*

*Statistical analysis: Nimesh Shah, Zhiruo Wan, Mao Hu, Meng Chen, Xinxin Lin, Jing Wang, Yue Wu Obtained funding: Yoganand Chillarige, Richard A. Forshee, Steven A. Anderson*

*Administrative, technical, or material support: Patricia C. Lloyd, Purva B. Shah, Nimesh Shah, Narayan Nair, Zhiruo Wan, Mao Hu, Tainya C. Clarke, Meng Chen, Xinxin Lin, Rose Do, Jing Wang, Yue Wu, Yoganand Chillarige, Richard A. Forshee, Steven A. Anderson*

*Supervision: Patricia C. Lloyd, Purva B. Shah, Mao Hu, Yoganand Chillarige, Richard A. Forshee, Steven A. Anderson*

### Data Access, Responsibility, and Analysis

Yoganand Chillarige had full access to all the data in the study and takes responsibility for the integrity of the data and the accuracy of the data analysis.

### Data Sharing Statement

Data from this study will not be shared.

## Acknowledgments

We thank Christine Hessler, Pengfei Zhang, Vivek Krishna, Thuy Nguyen, and Tyffany Chen for their support with the review of medical records of GBS cases. We thank Bradley Lufkin for his support with manuscript drafting and review. We thank Dr. Jane Gwira and Dr. Merianne Spencer for their review of the manuscript.

## Meeting Presentation

This work was presented at the Meeting of the Advisory Committee on Immunization Practices (ACIP); Atlanta, Georgia, on October 24, 2024 and at the 16^th^ Annual Sentinel Initiative Public Workshop; Washington, DC, on November 7, 2024.

## REFERENCES

1. Centers for Disease Control and Prevention (CDC). Respiratory Syncytial Virus (RSV) Infection. 11/7/2023; Available from: https://www.cdc.gov/rsv/index.html.

2. Melgar M, B.A., Roper LE, et al., Use of Respiratory Syncytial Virus Vaccines in Older Adults: Recommendations of the Advisory Committee on Immunization Practices — United States, 2023. Centers for Disease Control and Prevention (CDC): Morbidity and Mortality Weekly Report (MMWR).

3. United States (U.S.) Food and Drug Administration (FDA). FDA Approves First Respiratory Syncytial Virus (RSV) Vaccine 2023 May 04, 2023; Arexvy Approved for Individuals 60 Years of Age and Older]. Available from: https://www.fda.gov/news-events/press-announcements/fda-approves-first-respiratory-syncytial-virus-rsv-vaccine.

4. United States (U.S.) Food and Drug Administration (FDA). ABRYSVO. March 18, 2024; Available from: https://www.fda.gov/vaccines-blood-biologics/abrysvo.

5. United States Food and Drug Administration (FDA). Approval of mRESVIA (RSV Vaccine) for the prevention of LRTD caused by RSV in individuals 60 years of age and older. 2024 May 31, 2024; Available from: https://www.fda.gov/news-events/press-announcements/fda-roundup-may-31-2024.

6. United States (U.S.) Food and Drug Administration (FDA). AREXVY. 2024 08/22/2024; Available from: https://www.fda.gov/vaccines-blood-biologics/arexvy.

7. Britton A, R.L., Kotton CN, et al., Use of Respiratory Syncytial Virus Vaccines in Adults Aged ≥60 Years: Updated Recommendations of the Advisory Committee on Immunization Practices. 2024, Centers for Disease Control and Prevention: Morbidity and Mortality Weekly Report (MMWR).

8. United States (U.S.) Food and Drug Administration (FDA). Vaccines - Center for Biologics Evaluation and Research (CBER). January 12, 2024; Available from: https://www.fda.gov/vaccines-blood-biologics/vaccines.

9. United States (U.S.) Food and Drug Administration (FDA), AREXVY Package Insert - Highlights of Prescribing Information. 2023.

10. United States (U.S.) Food and Drug Administration (FDA), ABRYSVO Package Insert - Highlights of Prescribing Information. 2023.

11. Hause AM, M.P., Baggs J, et al., Early Safety Findings Among Persons Aged ≥60 Years Who Received a Respiratory Syncytial Virus Vaccine — United States.. 2024, Centers for Disease Control and Prevention (CDC): Morbidity and Mortality Weekly Report (MMWR).

12. Biologics Effectiveness and Safety (BEST) Initiative. Evaluation of Multiple Safety Outcomes following Respiratory Syncytial Virus (RSV) Vaccination in Adults 60 Years and Older December 22, 2022; Available from: https://bestinitiative.org/wp-content/uploads/2024/01/BEST_RSV_Safety_Older_Adults_2023-2024.pdf.

13. Petersen, I., I. Douglas, and H. Whitaker, Self controlled case series methods: an alternative to standard epidemiological study designs. 2016. 354: p. i4515.

14. Centers for Medicare & Medicaid Services (CMS). Minimum Data Set (MDS) 3.0 for Nursing Homes and Swing Bed Providers. September 10, 2024; Available from: https://www.cms.gov/medicare/quality/nursing-home-improvement/minimum-data-sets-swing-bed-providers.

15. Kind, A.J.H. and W.R. Buckingham, Making Neighborhood-Disadvantage Metrics Accessible - The Neighborhood Atlas. N Engl J Med, 2018. 378(26): p. 2456–2458.

16. Dodd, C.N., Romio, S. A., Black, S., et al., International collaboration to assess the risk of Guillain Barré Syndrome following Influenza A (H1N1) 2009 monovalent vaccines. Vaccine, 2013. 31(40): p. 4448–58.

17. Sejvar, J.J., Kohl, Katrin S., Gidudu, Jane, et al., Guillain–Barré syndrome and Fisher syndrome: Case definitions and guidelines for collection, analysis, and presentation of immunization safety data. Vaccine, 2011. 29(3): p. 599–612.

18. Rubin, D.B. and N. Schenker, Multiple Imputation for Interval Estimation From Simple Random Samples With Ignorable Nonresponse. Journal of the American Statistical Association, 1986. 81(394): p. 366–374.

19. Cox, C. and X. Li, Model-Based Estimation of the Attributable Risk: A Loglinear Approach. Comput Stat Data Anal, 2012. 56(12): p. 4180–4189.

20. Farrington, C.P., H.J. Whitaker, and M.N. Hocine, Case series analysis for censored, perturbed, or curtailed post-event exposures. Biostatistics, 2008. 10(1): p. 3–16.

21. United States (U.S.) Department of Health and Human Services (DHHS). 45 CFR 46 - Common Rule - Protections for Research Subjects. March 01, 2021 May 20, 2024]; Available from: https://www.hhs.gov/ohrp/regulations-and-policy/regulations/45-cfr-46/index.html.

22. Arya, D.P., Said, Maria A., Izurieta, Hector S., et al., Surveillance for Guillain-Barré syndrome after 2015–2016 and 2016–2017 influenza vaccination of Medicare beneficiaries. Vaccine, 2019. 37(43): p. 6543–6549.

23. Perez-Vilar, S., Hu, M., Weintraub, E., et al., Guillain-Barré Syndrome After High-Dose Influenza Vaccine Administration in the United States, 2018-2019 Season. J Infect Dis, 2021. 223(3): p. 416-425.

24. Goud, R., Lufkin, Bradley, Duffy, Jonathan, et al., Risk of Guillain-Barré Syndrome Following Recombinant Zoster Vaccine in Medicare Beneficiaries. JAMA Internal Medicine, 2021. 181(12): p. 1623–1630.

