## Supplemental File for "Evaluation of Guillain-Barré syndrome following Respiratory Syncytial Virus Vaccination among Medicare Beneficiaries 65 Years and Older"

| **List of Supplemental Tables and Figures** | **Supplemental Table and Figure Titles** |
| --- | --- |
| eTable1 | End-of-Season, Early-Season, and Observed vs. Expected Analyses: Summary of Analytical Plan and Study Specifications |
| eTable 2 | Secondary Analyses: IRR and AR with Corresponding 95% CI of GBS following RSV Vaccination for All Claims-Identified GBS Cases |
| eTable 3 | Secondary Analyses: IRR and AR with Corresponding 95% CI of GBS following RSV Vaccination for PPV-Based Imputation Analyses, and Chart-Confirmed GBS Cases |
| eTable 4 | Sensitivity Analyses: IRR and AR with Corresponding 95% CI of GBS following RSV Vaccination for PPV-Based Imputation Analyses, Chart-Confirmed, and All Claims-Identified GBS Cases |
| eTable 5 | Early-Season SCCS Analyses: IRR and AR with Corresponding 95% CI of GBS following RSV Vaccination for All Claims-Identified GBS Cases |
| eTable 6 | Observed vs. Expected Analysis: PPV-Based Imputation IRR and Cumulative Incidence with Corresponding 95% CI of GBS following RSV Vaccination |
| eFigure 1 | Hypothetical Example of Observation Period, Risk and Control Interval for Individual with a Qualifying GBS Outcome Following RSV Vaccination |
| eFigure 2 | Secondary Analyses: IRR with Corresponding 95% CI of GBS following RSV Vaccination for All Claims-Identified GBS Cases |
| eFigure 3 | Secondary Analyses: IRR with Corresponding 95% CI of GBS following RSV Vaccination for PPV-Based Imputation Analyses and Chart-Confirmed GBS Cases |
| eFigure 4 | Sensitivity Analyses: IRR with Corresponding 95% CI of GBS following RSV Vaccination for PPV-Based Imputation Analyses, Chart-Confirmed, and All Claims-Identified GBS Cases |
| eFigure 5 | Early-Season SCCS Analyses: IRR with Corresponding 95% CI of GBS following RSV Vaccination for All Claims-Identified GBS Cases |

**eTable 1: End-of-Season and Early-Season SCCS, and Observed vs. Expected Analyses: Summary of Analytical Plan and Study Specifications**

| **Analysis Specifications** | **End-of-Season SCCS Analysis** | | **Early-Season SCCS Analysis** | **Observed vs. Expected Analysis** |
| --- | --- | --- | --- | --- |
| Overview | The Respiratory Syncytial Virus (RSV) vaccines approved for use in adults 60 years of age and older in the United States (U.S.) are GlaxoSmithKline RSVPreF3+AS01 (AREXVY®), Pfizer RSVPreF (ABRYSVO®), and Moderna mRNA-1345 (mRESVIA®). RSVPreF3+AS01 and RSVPreF were further approved for use in individuals 50-59 and 18-59 years of age, respectively, who are at increased risk for Lower Respiratory Tract Disease (LRTD) caused by RSV.  This analysis used a self-controlled case series (SCCS) design and compared the incidence of Guillain-Barré syndrome (GBS) following RSV vaccines - RSVPreF3+AS01 and RSVPreF - within a post-vaccination risk interval with the incidence during all other parts of the observation period (i.e., control interval).  Only persons with an incident GBS outcome occurring in the study observation period (i.e., case population) are sampled in the SCCS study design. | | Consistent with End-of-Season SCCS analysis | This analysis used a retrospective cohort design with a historical comparator group.  It aimed to compare observed rates of GBS following RSV vaccines - RSVPreF3+AS01 and RSVPreF to the expected rates in the historical control population. |
| Data Sources | The study used administrative claims data from Centers for Medicare & Medicaid Services (CMS) Fee-for-Service (FFS) (Part A and B) from Medicare Shared Systems Data (SSD) and claims from Medicare Part D data.  Health service utilization for millions of enrollees was captured across multiple care settings – inpatient (IP), outpatient-emergency department (OP-ED), outpatient and professional (OP/PB), and pharmacy settings.  Beneficiaries’ demographic characteristics and information on death were captured from Medicare Enrollment Database (EDB).  Beneficiaries' nursing home residence information was assessed using assessment data from the Minimum Data Set (MDS).  Socioeconomic deprivation of beneficiary area of residence was assessed using the Area Deprivation Index (ADI). | | Consistent with End-of-Season SCCS analysis | Consistent with the SCCS analyses |
| Study Period | May 3, 2023 - July 13, 2024 | | May 3, 2023 - April 6, 2024 | May 3, 2023 - December 2, 2023 |
| Study Population | Eligible study population included Medicare beneficiaries 65 years of age and older enrolled in Medicare FFS and Medicare Part D on the date of their first observed RSV vaccination, who received an RSV vaccine during the study period and were not missing age and sex information.  For inclusion in the GBS outcome-specific analyses, beneficiaries were required to:   - Be continuously enrolled in Medicare FFS from 365 days prior to RSV vaccination, - Receive a single dose of either of the RSV vaccine product prior to the vaccination data cutoff date (**January 28, 2024**), and - Experience an incident GBS outcome during the study period, with no prior outcome in the GBS outcome-specific clean window relative to the outcome date | | Eligible study population included Medicare beneficiaries 65 years of age and older enrolled in Medicare FFS and Medicare Part D on the date of their first observed RSV vaccination, who received an RSV vaccine during the study period and were not missing age and sex information.  For inclusion in the GBS outcome-specific analyses, beneficiaries were required to:   - Be continuously enrolled in Medicare FFS from 365 days prior to RSV vaccination, - Receive a single dose of either of the RSV vaccine product prior to the vaccination data cutoff date (**October 22, 2023**), and - Experience an incident GBS outcome during the study period, with no prior outcome in the GBS outcome-specific clean window relative to the outcome date | Eligible study population included Medicare beneficiaries 65 years of age and older enrolled in Medicare FFS and Medicare Part D on the date of their first observed RSV vaccination, who received an RSV vaccine during the study period and were not missing age and sex information.  For inclusion in the GBS outcome-specific analyses, beneficiaries were required to:   - Be continuously enrolled in Medicare FFS from the GBS-specific clean window to their first observed RSV vaccination, - Receive a single dose of either of the RSV vaccine product within the study period (May 3 [RSVPreF3+AS01] or May 31, 2023 [RSVPreF3] – **December 2, 2023**), and |
| Exposure | Receipt of RSVPreF3+AS01 and RSVPreF vaccines was identified using brand-specific Current Procedural Terminology (CPT)/ Healthcare Common Procedure Coding System (HCPCS) codes or National Drug Codes (NDCs) in prescription drug or other care settings. | | Consistent with End-of-Season SCCS analysis | Consistent with the SCCS analyses |
|  | RSVPreF3+AS01:  HCPCS/CPT: 90679  NDC:  58160072303, 58160074403, 58160084811 | RSVPreF:  HCPCS/CPT: 90678  NDC:  00069020701, 00069025001, 00069034401, 00069034405,  00069034410 |  |  |
| Health Outcome | Incident occurrences of GBS were identified using International Classification of Disease, 10th Revision, Clinical Modification (ICD-10-CM) diagnosis codes.  The first occurrence of GBS outcome was defined as an incident case if no outcome was recorded during the preceding clean window relative to the outcome date.  GBS: ICD-10-CM diagnosis code G610 in the primary diagnosis position on IP claims Clean Window: 365 days Risk Interval: 1-42 days Control Interval: 43-90 days | | Consistent with End-of-Season SCCS analysis | Incident occurrences of GBS were identified using ICD-10-CM diagnosis codes. The first occurrence of GBS outcome was defined as an incident case if no outcome was recorded during the preceding the clean window relative to the exposure date.  GBS: ICD-10-CM diagnosis code G610 in the primary diagnosis position on IP claims Clean Window: 365 days Risk Interval: 1-42 days |
| Observation Period | The observation period extends from the RSV vaccination date through 90 days after vaccination with either RSV vaccine.  Beneficiaries are censored at the first of disenrollment, death, or the end of the study period. The vaccination cutoff date (i.e., **January 28, 2024)**, was set to ensure at least 90% data completeness at the end of the observation period to ensure complete capture of GBS outcome | | The observation period extends from the RSV vaccination date through 90 days after vaccination with either RSV vaccine.  Beneficiaries are censored at the first of disenrollment, death, or study period end. The vaccination cutoff date (i.e., **October 22, 2023**), was set to ensure at least 90% data completeness at the end of observation period to ensure complete capture of GBS outcome | The observation period extends from the RSV vaccination date through **December 2, 2023**. Beneficiaries are censored at the first of outcome occurrence, death, disenrollment, end of the outcome-specific risk interval, subsequent RSV vaccination, or end of the study period. |
| Risk and Control Intervals | A post-vaccination risk interval is defined as the time during which excess risk is hypothesized following one dose of either of the RSV vaccines for the outcome based on biological plausibility and clinical input.  The post-vaccination control interval is defined as all follow-up time during the observation period following one dose of either of the RSV vaccines that is outside of the risk interval. | | Consistent with End-of-Season SCCS analysis | A post-vaccination risk interval is defined as the time during which excess risk is hypothesized following RSV vaccination for the outcome based on biological plausibility and clinical input. |
| Medical Record Review (MRR) | A MRR was conducted on all GBS cases identified in Medicare claims that qualified for the end-of-season SCCS analysis (including GBS cases identified in early-season SCCS analysis).  Medical records were requested and each record was adjudicated by two independent neurologists using the Brighton Collaboration's case definition for GBS, where cases classified as Brighton Levels 1, 2, or 3 of diagnostic certainty were treated as "chart-confirmed" GBS cases.  Positive predictive values (PPVs) with 95% confidence intervals (CIs) were calculated as the percentage of received records classified as chart-confirmed through the adjudication process. | | All GBS cases identified in the Medicare claims for the early-season SCCS analysis were included in the MRR for the end-of-season SCCS analysis | All GBS cases identified in the Medicare claims for the observed vs. expected analysis were included in the MRR for the end-of-season SCCS analysis |
| Statistical Analyses | Inferential analyses were conducted on:   - All claims-identified GBS cases, - Chart-confirmed GBS cases, and - Chart-confirmed and unreturned cases with a PPV-based imputation on unreturned cases (i.e., PPV-based imputation analyses cases)   Incidence Rate Ratio (IRR), the ratio of the incidence in the exposure risk intervals relative to the incidence in control intervals for GBS, were estimated using conditional Poisson regression model.  Attributable risk (AR) was estimated as the number of excess GBS cases per 100,000 doses or person-years following RSV vaccination compared to not receiving an RSV vaccine.  Statistical significance was determined using two-sided hypothesis tests with an alpha of 0.05. | | Inferential analyses were conducted only on:   - All claims-identified GBS cases since the MRR of GBS following RSV vaccination was in progress at the time of the early-season SCCS analysis   IRR, the ratio of the incidence in the exposure risk intervals relative to the incidence in control intervals for GBS, were estimated using conditional Poisson regression model.  AR was estimated as the number of excess GBS cases per 100,000 doses or person-years following RSV vaccination compared to not receiving an RSV vaccine.  Statistical significance was determined using two-sided hypothesis tests with an alpha of 0.05. | Historical GBS comparator rates were calculated from the 2022 calendar year.  For each RSV vaccine product, the observed incidence rates (IRs) per 100,000 person-years were calculated for GBS and the corresponding 95% CIs were estimated based on a Poisson model.  The observed IRs were divided by the historical comparator rates, to obtain IRRs for GBS.  The observed rates and IRRs for GBS were assessed by sex, age group (65-74 years, 75-84 years, 85 years and older), and any concomitant vaccine administration when sufficient counts were available in the historical comparator population (data not shown). |
| Adjustment to the Statistical Analyses to Control Confounding | Inferential analyses conducted on all claims-identified and chart-confirmed GBS cases, included adjustments for:   - Outcome-dependent observation time (i.e., censoring of observation period due to death and disenrollment), also referred to as the Farrington adjustment, and - Seasonality adjustment using incidence rates of GBS estimated from Medicare FFS population aged 65 years and older from corresponding calendar months in 2022-2023 used as baseline incidence of GBS in risk and control intervals   Inferential analyses conducted on PPV-based imputation analyses cases included adjustment for:   - PPV-based imputation on unreturned cases utilized a quantitative bias analysis, where multiple datasets (10,000) were created where the status of those non-returned cases were imputed by assigning them the status of chart-confirmed with the probability equal to PPV derived from the MRR of GBS following RSV vaccination. A conditional Poisson regression was repeated on multiple datasets and the IRR estimates were pooled across these datasets. | | Inferential analyses conducted on all claims identified GBS cases, included adjustments for:   - Outcome-dependent observation time (i.e., censoring of observation period due to death and disenrollment), also referred to as the Farrington adjustment, - Seasonality adjustment using incidence rates of GBS estimated from Medicare FFS population aged 65 years and older from corresponding calendar months in 2022-2023 used as baseline incidence of GBS in risk and control intervals, and - PPV-based imputation adjustment using quantitative bias analysis by utilizing the PPV available from a prior medical record review (MRR) of GBS following influenza vaccination (PPV: 71.21%), since the MRR of GBS following RSV vaccination was in progress at the time of the early-season SCCS analysis | The observed vs. expected analysis included adjustment for:   - PPV using a quantitative bias analysis by utilizing the PPV available from a prior medical record review (MRR) of GBS following influenza vaccination (PPV: 71.21%), since the MRR of GBS following RSV vaccination was in progress at the time of the early season SCCS analysis |
| Additional Analyses to Control Confounding | Secondary analyses were conducted for each RSV vaccine product with a statistically significant (alpha = 0.1) elevation of IRR in the primary SCCS analyses, to account for potential effect of same-day concomitant vaccination with RSV vaccination. IRRs and ARs with their corresponding 95% CIs were calculated for GBS among the study population with and without concomitant vaccines on the same day as their RSV vaccination. Concomitant vaccination was defined as receipt of 2023-2024 influenza or Coronavirus Disease 2019 (COVID-19) vaccine, pneumococcal conjugate vaccine (PCV), or Shingles vaccines, on the same date as the RSV vaccination, each identified using CPT/ HCPCS or NDCs on administrative claims data.  Sensitivity analyses were conducted that:   - Incorporated a 14-day washout interval between risk and control intervals to evaluate potential bias associated with carryover effects from RSV vaccination contributing to the risk in control intervals, and - Included an individual’s full observation period length for persons that died or disenrolled to evaluate potential bias from a violation of the SCCS assumption that observation length is independent of outcomes | | Consistent with End-of-Season SCCS analysis (data not shown) | N/A |

**eTable 2: Secondary Analyses: IRR and AR with Corresponding 95% CIs of GBS following RSV Vaccination for All Claims-Identified GBS Cases**

| **Secondary Analyses by RSV Vaccine Product** | **Eligible Vaccines** | **Cases in Risk Interval** | **Cases in Control Interval** | **Model Estimate** | | **AR Per 100,000 Vaccine Doses**  **(95% CI)** | **AR Per 100,000 Person-Years**  **(95% CI)** |
| --- | --- | --- | --- | --- | --- | --- | --- |
|  |  |  |  | **IRR with 95% CI** | **P-Value ^a^** |  |  |
| ***Seasonality and Farrington Adjustment*** | | | | | | | |
| RSVPreF3+AS01 *With*  Concomitant Vaccination | 833,067 | ** | <11 | 2.19 (0.87, 5.49) | 0.09 | 0.85 (-0.09, 1.79) | 7.40 (-0.79, 15.59) |
| RSVPreF3+AS01 *Without*  Concomitant Vaccination | 1,369,180 | ** | <11 | 3.47 (1.61, 7.46) | **<0.001** | 1.40 (0.72, 2.09) | 12.27 (6.26, 18.28) |
| RSVPreF *With*  Concomitant Vaccination | 420,764 | ** | <11 | 2.26 (0.89, 5.73) | 0.09 | 1.59 (-0.18, 3.35) | 13.85 (-1.55, 29.25) |
| RSVPreF *Without*  Concomitant Vaccination | 603,678 | ** | <11 | 4.48 (1.50, 13.42) | **0.01** | 2.06 (0.99, 3.12) | 18.01 (8.70, 27.31) |
| ***Farrington Adjustment*** | | | | | | | |
| RSVPreF3+AS01 *With*  Concomitant Vaccination | 833,067 | ** | <11 | 2.13 (0.85, 5.34) | 0.11 | 0.83 (-0.13, 1.79) | 7.22 (-1.14, 15.58) |
| RSVPreF3+AS01 *Without*  Concomitant Vaccination | 1,369,180 | ** | <11 | 3.58 (1.67, 7.70) | **<0.001** | 1.42 (0.75, 2.10) | 12.43 (6.53, 18.33) |
| RSVPreF *With*  Concomitant Vaccination | 420,764 | ** | <11 | 2.04 (0.80, 5.19) | 0.13 | 1.46 (-0.37, 3.29) | 12.71 (-3.25, 28.68) |
| RSVPreF *Without*  Concomitant Vaccination | 603,678 | ** | <11 | 4.59 (1.53, 13.72) | **0.01** | 2.07 (1.03, 3.12) | 18.13 (9.00, 27.26) |
| ***Seasonality Adjustment*** | | | | | | | |
| RSVPreF3+AS01 *With*  Concomitant Vaccination | 833,067 | ** | <11 | 2.18 (0.87, 5.48) | 0.10 | 0.85 (-0.09, 1.79) | 7.38 (-0.82, 15.58) |
| RSVPreF3+AS01 *Without*  Concomitant Vaccination | 1,369,180 | ** | <11 | 3.19 (1.49, 6.80) | **<0.001** | 1.35 (0.64, 2.07) | 11.83 (5.58, 18.07) |
| RSVPreF *With*  Concomitant Vaccination | 420,764 | ** | <11 | 2.06 (0.81, 5.25) | 0.13 | 1.47 (-0.35, 3.28) | 12.79 (-3.02, 28.59) |
| RSVPreF *Without*  Concomitant Vaccination | 603,678 | ** | <11 | 4.47 (1.49, 13.38) | **0.01** | 2.06 (0.99, 3.12) | 17.99 (8.67, 27.31) |
| ***Non-Farrington and Non-Seasonality Adjustment*** | | | | | | | |
| RSVPreF3+AS01 *With*  Concomitant Vaccination | 833,067 | ** | <11 | 2.12 (0.85, 5.32) | 0.11 | 0.83 (-0.13, 1.78) | 7.20 (-1.17, 15.58) |
| RSVPreF3+AS01 *Without*  Concomitant Vaccination | 1,369,180 | ** | <11 | 3.29 (1.54, 7.02) | **<0.001** | 1.37 (0.67, 2.07) | 12.00 (5.88, 18.13) |
| RSVPreF *With*  Concomitant Vaccination | 420,764 | ** | <11 | 1.86 (0.73, 4.76) | 0.19 | 1.32 (-0.56, 3.20) | 11.54 (-4.86, 27.94) |
| RSVPreF *Without*  Concomitant Vaccination | 603,678 | ** | <11 | 4.57 (1.53, 13.67) | **0.01** | 2.07 (1.03, 3.12) | 18.11 (8.97, 27.25) |

^a^ p<0.05 are highlighted in bold

AR = Attributable Risk, CI = Confidence Interval, GBS = Guillain-Barré Syndrome, IRR = Incidence Rate Ratio, RSV = Respiratory Syncytial Virus

Cell sizes <11 are suppressed to protect patient confidentiality; **Display of specific cell counts discloses small cell sizes <11

**eTable 3: Secondary Analyses: IRR and AR with Corresponding 95% CIs of GBS following RSV Vaccination for PPV-Based Imputation Analyses, and Chart-Confirmed GBS Cases**

| **Secondary Analyses by RSV Vaccine Product** | **Eligible Vaccines** | **Cases in Risk Interval** | **Cases in Control Interval** | **Model Estimate** | | **AR Per 100,000 Vaccine Doses**  **(95% CI)** | **AR Per 100,000 Person-Years**  **(95% CI)** |
| --- | --- | --- | --- | --- | --- | --- | --- |
|  |  |  |  | **IRR with 95% CI** | **P-Value ^a^** |  |  |
| **PPV-Based Imputation Analyses Cases (n=71) ^b^** | | | | | | | |
| ***PPV-Based Imputation, Seasonality, and Farrington Adjustment – [Risk and Control Interval PPVs Applied to Respective Unreturned Cases]*** | | | | | | | |
| RSVPreF3+AS01 *With*  Concomitant Vaccination | 833,067 | <11 | <11 | 2.45 (0.69, 8.71) | 0.17 | 0.56 (-0.19, 1.31) | 4.88 (-1.69, 11.44) |
| RSVPreF3+AS01 *Without*  Concomitant Vaccination | 1,369,180 | 16 | <11 | 2.46 (1.01, 5.98) | 0.05 | 0.71 (0.07, 1.35) | 6.20 (0.65, 11.76) |
| RSVPreF *With*  Concomitant Vaccination | 420,764 | <11 | <11 | 1.49 (0.51, 4.39) | 0.47 | 0.62 (-1.08, 2.32) | 5.38 (-9.46, 20.21) |
| RSVPreF *Without*  Concomitant Vaccination | 603,678 | <11 | <11 | 3.19 (0.90, 11.29) | 0.07 | 1.17 (0.16, 2.18) | 10.20 (1.38, 19.02) |
| ***PPV-Based Imputation, Seasonality, and Farrington Adjustment – [Overall PPV Applied to Unreturned Cases]*** | | | | | | | |
| RSVPreF3+AS01 *With*  Concomitant Vaccination | 833,067 | <11 | <11 | 2.61 (0.72, 9.44) | 0.14 | 0.59 (-0.16, 1.35) | 5.19 (-1.38, 11.76) |
| RSVPreF3+AS01 *Without*  Concomitant Vaccination | 1,369,180 | 17 | <11 | 2.62 (1.06, 6.44) | **0.04** | 0.76 (0.12, 1.39) | 6.62 (1.06, 12.17) |
| RSVPreF *With*  Concomitant Vaccination | 420,764 | <11 | <11 | 1.53 (0.53, 4.45) | 0.44 | 0.66 (-1.04, 2.36) | 5.78 (-9.06, 20.61) |
| RSVPreF *Without*  Concomitant Vaccination | 603,678 | <11 | <11 | 3.54 (0.93, 13.43) | 0.06 | 1.23 (0.22, 2.24) | 10.78 (1.96, 19.60) |
| ***PPV-Based Imputation and Seasonality Adjustment – [Overall PPV Applied to Unreturned Cases]*** | | | | | | | |
| RSVPreF3+AS01 *With*  Concomitant Vaccination | 833,067 | <11 | <11 | 2.60 (0.72, 9.41) | 0.15 | 0.59 (-0.16, 1.35) | 5.18 (-1.40, 11.76) |
| RSVPreF3+AS01 *Without*  Concomitant Vaccination | 1,369,180 | 17 | <11 | 2.51 (1.02, 6.18) | **0.04** | 0.74 (0.09, 1.39) | 6.45 (0.75, 12.15) |
| RSVPreF *With*  Concomitant Vaccination | 420,764 | <11 | <11 | 1.38 (0.49, 3.88) | 0.55 | 0.52 (-1.22, 2.26) | 4.55 (-10.63, 19.73) |
| RSVPreF *Without*  Concomitant Vaccination | 603,678 | <11 | <11 | 3.53 (0.93, 13.39) | 0.06 | 1.23 (0.22, 2.24) | 10.77 (1.93, 19.60) |
| ***PPV-Based Imputation and Farrington Adjustment – [Overall PPV Applied to Unreturned Cases]*** | | | | | | | |
| RSVPreF3+AS01 *With*  Concomitant Vaccination | 833,067 | <11 | <11 | 2.51 (0.69, 9.09) | 0.16 | 0.58 (-0.18, 1.35) | 5.07 (-1.61, 11.75) |
| RSVPreF3+AS01 *Without*  Concomitant Vaccination | 1,369,180 | 17 | <11 | 2.70 (1.10, 6.65) | **0.03** | 0.77 (0.15, 1.39) | 6.75 (1.30, 12.19) |
| RSVPreF *With*  Concomitant Vaccination | 420,764 | <11 | <11 | 1.39 (0.48, 4.04) | 0.55 | 0.53 (-1.23, 2.30) | 4.67 (-10.71, 20.04) |
| RSVPreF *Without*  Concomitant Vaccination | 603,678 | <11 | <11 | 3.62 (0.96, 13.71) | 0.06 | 1.24 (0.26, 2.23) | 10.88 (2.27, 19.49) |
| ***PPV-Based Imputation Adjustment – [Overall PPV Applied to Unreturned Cases]*** | | | | | | | |
| RSVPreF3+AS01 *With*  Concomitant Vaccination | 833,067 | <11 | <11 | 2.50 (0.69, 9.06) | 0.16 | 0.58 (-0.19, 1.35) | 5.06 (-1.63, 11.74) |
| RSVPreF3+AS01 *Without*  Concomitant Vaccination | 1,369,180 | 17 | <11 | 2.60 (1.06, 6.39) | **0.04** | 0.75 (0.11, 1.39) | 6.58 (1.00, 12.17) |
| RSVPreF *With*  Concomitant Vaccination | 420,764 | <11 | <11 | 1.25 (0.44, 3.52) | 0.68 | 0.38 (-1.42, 2.18) | 3.31 (-12.43, 19.06) |
| RSVPreF *Without*  Concomitant Vaccination | 603,678 | <11 | <11 | 3.61 (0.95, 13.67) | 0.06 | 1.24 (0.26, 2.23) | 10.87 (2.24, 19.49) |
| **Chart-Confirmed Cases (n=51)** | | | | | | | |
| ***Seasonality and Farrington Adjustment*** | | | | | | | |
| RSVPreF3+AS01 *With*  Concomitant Vaccination | 833,067 | <11 | <11 | 2.44 (0.61, 9.77) | 0.21 | 0.43 (-0.14, 0.99) | 3.71 (-1.26, 8.68) |
| RSVPreF3+AS01 *Without*  Concomitant Vaccination | 1,369,180 | ** | <11 | 2.35 (0.88, 6.27) | 0.09 | 0.50 (-0.04, 1.04) | 4.40 (-0.32, 9.13) |
| RSVPreF *With*  Concomitant Vaccination | 420,764 | <11 | <11 | 1.09 (0.36, 3.23) | 0.88 | 0.11 (-1.36, 1.58) | 0.98 (-11.85, 13.82) |
| RSVPreF *Without*  Concomitant Vaccination | 603,678 | <11 | <11 | 5.15 (1.11, 23.87) | **0.04** | 1.20 (0.50, 1.91) | 10.51 (4.33, 16.69) |
| ***Seasonality Adjustment*** | | | | | | | |
| RSVPreF3+AS01 *With*  Concomitant Vaccination | 833,067 | <11 | <11 | 2.43 (0.61, 9.74) | 0.21 | 0.42 (-0.15, 0.99) | 3.70 (-1.27, 8.68) |
| RSVPreF3+AS01 *Without*  Concomitant Vaccination | 1,369,180 | ** | <11 | 2.24 (0.84, 5.96) | 0.11 | 0.48 (-0.08, 1.05) | 4.24 (-0.68, 9.15) |
| RSVPreF *With*  Concomitant Vaccination | 420,764 | <11 | <11 | 1.08 (0.36, 3.22) | 0.89 | 0.11 (-1.36, 1.58) | 0.95 (-11.90, 13.80) |
| RSVPreF *Without*  Concomitant Vaccination | 603,678 | <11 | <11 | 5.14 (1.11, 23.79) | **0.04** | 1.20 (0.49, 1.91) | 10.50 (4.32, 16.69) |
| ***Farrington Adjustment*** | | | | | | | |
| RSVPreF3+AS01 *With*  Concomitant Vaccination | 833,067 | <11 | <11 | 2.29 (0.57, 9.17) | 0.24 | 0.41 (-0.18, 0.99) | 3.54 (-1.59, 8.68) |
| RSVPreF3+AS01 *Without*  Concomitant Vaccination | 1,369,180 | ** | <11 | 2.39 (0.90, 6.37) | 0.08 | 0.51 (-0.02, 1.04) | 4.46 (-0.20, 9.11) |
| RSVPreF *With*  Concomitant Vaccination | 420,764 | <11 | <11 | 0.98 (0.33, 2.92) | 0.97 | -0.03 (-1.54, 1.49) | -0.22 (-13.45, 13.01) |
| RSVPreF *Without*  Concomitant Vaccination | 603,678 | <11 | <11 | 5.16 (1.11, 23.87) | **0.04** | 1.20 (0.50, 1.90) | 10.51 (4.42, 16.61) |
| ***Non-Farrington and Non-Seasonality Adjustment*** | | | | | | | |
| RSVPreF3+AS01 *With*  Concomitant Vaccination | 833,067 | <11 | <11 | 2.29 (0.57, 9.14) | 0.24 | 0.41 (-0.18, 0.99) | 3.54 (-1.61, 8.68) |
| RSVPreF3+AS01 *Without*  Concomitant Vaccination | 1,369,180 | ** | <11 | 2.27 (0.85, 6.05) | 0.10 | 0.49 (-0.06, 1.04) | 4.29 (-0.55, 9.13) |
| RSVPreF *With*  Concomitant Vaccination | 420,764 | <11 | <11 | 0.98 (0.33, 2.91) | 0.97 | -0.03 (-1.55, 1.49) | -0.26 (-13.51, 12.99) |
| RSVPreF *Without*  Concomitant Vaccination | 603,678 | <11 | <11 | 5.14 (1.11, 23.80) | **0.04** | 1.20 (0.50, 1.90) | 10.50 (4.40, 16.61) |

^a^ p<0.05 are highlighted in bold

^b^ Cases in the risk and control interval for PPV-based imputation analyses are the average number of true cases in the multiple imputation process; rounded to the nearest whole number

AR = Attributable Risk, CI = Confidence Interval, GBS = Guillain-Barré Syndrome, IRR = Incidence Rate Ratio, PPV = Positive Predictive Value, RSV = Respiratory Syncytial Virus

Cell sizes <11 are suppressed to protect patient confidentiality; **Display of specific cell counts discloses small cell sizes <11

**eTable 4: Sensitivity Analyses: IRR and AR with Corresponding 95% CIs of GBS following RSV Vaccination for PPV-Based Imputation Analyses, Chart-Confirmed, and All Claims-Identified GBS Cases**

| **Sensitivity Analyses by RSV Vaccine Product** | **Eligible Vaccines** | **Cases in Risk Interval** | **Cases in Control Interval** | **Model Estimate** | | **AR Per 100,000 Vaccine Doses**  **(95% CI)** | **AR Per 100,000 Person-Years**  **(95% CI)** |
| --- | --- | --- | --- | --- | --- | --- | --- |
|  |  |  |  | **IRR with 95% CI** | **P-Value ^a^** |  |  |
| **PPV-Based Imputation Analyses Cases (n=71) ^b^** | | | | | | | |
| **Assessment of Washout Period Between Risk and Control Intervals Analyses** | | | | | | | |
| *PPV-Based Imputation, Seasonality, and Farrington Adjustment - [Risk and Control Interval PPVs Applied to Respective Unreturned Cases]* | | | | | | | |
| RSVPreF3+AS01 | 2,202,247 | 24 | <11 | 2.03 (0.93, 4.42) | 0.07 | 0.56 (0.01, 1.11) | 4.88 (0.06, 9.70) |
| RSVPreF | 1,024,442 | 18 | <11 | 1.54 (0.69, 3.44) | 0.30 | 0.62 (-0.50, 1.73) | 5.39 (-4.35, 15.14) |
| **Full-Planned Observation Time Analyses** | | | | | | | |
| *PPV-Based Imputation and Seasonality Adjustment - [Risk and Control Interval PPVs Applied to Respective Unreturned Cases]* | | | | | | | |
| RSVPreF3+AS01 | 2,202,247 | 24 | 11 | 2.40 (1.16, 4.95) | **0.02** | 0.64 (0.16, 1.12) | 5.61 (1.44, 9.78) |
| RSVPreF | 1,024,442 | 18 | <11 | 2.02 (0.93, 4.39) | 0.08 | 0.89 (-0.02, 1.81) | 7.81 (-0.22, 15.84) |
| *PPV-Based Imputation and Seasonality Adjustment - [Overall PPV Applied to Unreturned Cases]* | | | | | | | |
| RSVPreF3+AS01 | 2,202,247 | 25 | 11 | 2.55 (1.22, 5.31) | **0.01** | 0.68 (0.20, 1.17) | 5.98 (1.76, 10.20) |
| RSVPreF | 1,024,442 | 18 | <11 | 2.11 (0.97, 4.63) | 0.06 | 0.95 (-0.01, 1.91) | 8.28 (-0.11, 16.66) |
| *PPV-Based Imputation Adjustment - [Overall PPV Applied to Unreturned Cases]* | | | | | | | |
| RSVPreF3+AS01 | 2,202,247 | 25 | 11 | 2.57 (1.23, 5.37) | **0.01** | 0.69 (0.21, 1.17) | 6.02 (1.82, 10.21) |
| RSVPreF | 1,024,442 | 18 | <11 | 2.03 (0.93, 4.44) | 0.08 | 0.91 (-0.06, 1.88) | 7.97 (-0.51, 16.46) |
| **Chart-Confirmed Cases (n=51)** | | | | | | | |
| **Full-Planned Observation Time Analyses** | | | | | | | |
| *Seasonality Adjustment* | | | | | | | |
| RSVPreF3+AS01 | 2,202,247 | ** | <11 | 2.31 (1.04, 5.14) | **0.04** | 0.46 (0.07, 0.86) | 4.05 (0.58, 7.52) |
| RSVPreF | 1,024,442 | ** | <11 | 2.01 (0.88, 4.60) | 0.10 | 0.74 (-0.08, 1.56) | 6.43 (-0.74, 13.60) |
| *Non-Farrington and Non-Seasonality Adjustment* | | | | | | | |
| RSVPreF3+AS01 | 2,202,247 | ** | <11 | 2.29 (1.03, 5.09) | **0.04** | 0.46 (0.06, 0.86) | 4.02 (0.55, 7.48) |
| RSVPreF | 1,024,442 | ** | <11 | 1.90 (0.83, 4.35) | 0.13 | 0.70 (-0.14, 1.53) | 6.08 (-1.25, 13.40) |
| **All Claims-Identified Cases (n=95)** | | | | | | | |
| **Full-Planned Observation Time Analyses** | | | | | | | |
| *Seasonality Adjustment* | | | | | | | |
| RSVPreF3+AS01 | 2,202,247 | 40 | 16 | 2.83 (1.58, 5.05) | **<0.001** | 1.17 (0.61, 1.74) | 10.25 (5.29, 15.22) |
| RSVPreF | 1,024,442 | 28 | 11 | 3.02 (1.50, 6.07) | **<0.001** | 1.83 (0.85, 2.81) | 15.97 (7.41, 24.53) |
| *Non-Farrington and Non-Seasonality Adjustment* | | | | | | | |
| RSVPreF3+AS01 | 2,202,247 | 40 | 16 | 2.86 (1.60, 5.10) | **<0.001** | 1.18 (0.62, 1.75) | 10.32 (5.38, 15.25) |
| RSVPreF | 1,024,442 | 28 | 11 | 2.91 (1.45, 5.84) | **<0.001** | 1.79 (0.80, 2.79) | 15.67 (7.00, 24.34) |

^a^ p<0.05 are highlighted in bold

^b^ Cases in the risk and control interval for PPV-based imputation analyses are the average number of true cases in the multiple imputation process; rounded to the nearest whole number

AR = Attributable Risk, CI = Confidence Interval, GBS = Guillain-Barré Syndrome, IRR = Incidence Rate Ratio, PPV = Positive Predictive Value, RSV = Respiratory Syncytial Virus

Cell sizes <11 are suppressed to protect patient confidentiality; **Display of specific cell counts discloses small cell sizes <11

**eTable 5: Early-Season SCCS Analysis: IRR and AR with Corresponding 95% CIs of GBS following RSV Vaccination for All Claims-Identified GBS Cases**

| **Early-Season SCCS Analysis by RSV Vaccine Product** | **Eligible Vaccines** | **Cases in Risk Interval** | **Cases in Control Interval** | **Model Estimate** | | **AR Per 100,000 Vaccine Doses**  **(95% CI)** | **AR Per 100,000 Person-Years**  **(95% CI)** |
| --- | --- | --- | --- | --- | --- | --- | --- |
|  |  |  |  | **IRR with 95% CI** | **P-Value ^a^** |  |  |
| **All Claims-Identified Cases (n=28)** | | | | | | | |
| *PPV-Based Imputation, Seasonality, and Farrington Adjustment ^b^* | | | | | | | |
| RSVPreF3+AS01 | 872,068 | <11 | <11 | 2.30 (0.39, 13.72) | 0.36 | 0.32 (-0.30, 0.95) | 2.81 (-2.64, 8.26) |
| RSVPreF | 456,107 | <11 | <11 | 4.48 (0.88, 22.90) | 0.07 | 1.57 (0.30, 2.85) | 13.69 (2.59, 24.79) |
| *PPV-Based Imputation and Farrington Adjustment ^b^* | | | | | | | |
| RSVPreF3+AS01 | 872,068 | <11 | <11 | 2.04 (0.34, 12.11) | 0.43 | 0.29 (-0.37, 0.95) | 2.53 (-3.23, 8.29) |
| RSVPreF | 456,107 | <11 | <11 | 3.96 (0.77, 20.28) | 0.10 | 1.51 (0.15, 2.88) | 13.17 (1.27, 25.06) |
| *Seasonality Adjustment and Farrington Adjustment* | | | | | | | |
| RSVPreF3+AS01 | 872,068 | <11 | <11 | 2.27 (0.66, 7.75) | 0.19 | 0.45 (-0.18, 1.08) | 3.91 (-1.54, 9.36) |
| RSVPreF | 456,107 | ** | <11 | 4.27 (1.39, 13.14) | **0.01** | 2.18 (0.91, 3.46) | 19.00 (7.89, 30.10) |
| *Farrington Adjustment* | | | | | | | |
| RSVPreF3+AS01 | 872,068 | <11 | <11 | 2.01 (0.59, 6.85) | 0.27 | 0.40 (-0.26, 1.06) | 3.50 (-2.24, 9.25) |
| RSVPreF | 456,107 | ** | <11 | 3.71 (1.21, 11.42) | **0.02** | 2.08 (0.73, 3.43) | 18.12 (6.38, 29.87) |
| *PPV-Based Imputation and Seasonality Adjustment ^b^* | | | | | | | |
| RSVPreF3+AS01 | 872,068 | <11 | <11 | 2.30 (0.39, 13.68) | 0.36 | 0.32 (-0.30, 0.95) | 2.80 (-2.65, 8.26) |
| RSVPreF | 456,107 | <11 | <11 | 4.21 (0.83, 21.45) | 0.08 | 1.54 (0.24, 2.85) | 13.44 (2.08, 24.80) |
| *PPV-Based Imputation Adjustment ^b^* | | | | | | | |
| RSVPreF3+AS01 | 872,068 | <11 | <11 | 2.07 (0.35, 12.32) | 0.42 | 0.30 (-0.36, 0.95) | 2.57 (-3.15, 8.29) |
| RSVPreF | 456,107 | <11 | <11 | 3.74 (0.74, 19.01) | 0.11 | 1.48 (0.09, 2.88) | 12.91 (0.77, 25.06) |
| *Seasonality Adjustment* | | | | | | | |
| RSVPreF3+AS01 | 872,068 | <11 | <11 | 2.26 (0.66, 7.73) | 0.19 | 0.45 (-0.18, 1.07) | 3.90 (-1.56, 9.36) |
| RSVPreF | 456,107 | ** | <11 | 4.07 (1.32, 12.53) | **0.01** | 2.15 (0.84, 3.45) | 18.71 (7.35, 30.07) |
| *Non-Farrington Adjustment* | | | | | | | |
| RSVPreF3+AS01 | 872,068 | <11 | <11 | 2.04 (0.60, 6.97) | 0.25 | 0.41 (-0.24, 1.06) | 3.56 (-2.13, 9.26) |
| RSVPreF | 456,107 | ** | <11 | 3.61 (1.17, 11.13) | **0.03** | 2.06 (0.69, 3.43) | 17.94 (6.00, 29.89) |

^a^ p<0.05 are highlighted in bold

^b^ Cases in the risk and control interval for PPV-based imputation analyses are the average number of true cases in the multiple imputation process

AR = Attributable Risk, CI = Confidence Interval, GBS = Guillain-Barré Syndrome, IRR = Incidence Rate Ratio, PPV = Positive Predictive Value, RSV = Respiratory Syncytial Virus, SCCS = Self-Controlled Case Series

Cell sizes <11 are suppressed to protect patient confidentiality; **Display of specific cell counts discloses small cell sizes <11

**eTable 6: Observed vs. Expected Analysis: PPV-Based Imputation IRR and Cumulative Incidence with Corresponding 95% CI of GBS following RSV Vaccination**

| **RSV Vaccine Product** | **Age Category** | **Eligible Vaccines** | **Observed Outcomes** | **PPV-Based Imputed IRR (95% CI) ^a^** | **Cumulative Incidence Per 100,000 Vaccine Doses (95% CI)** |
| --- | --- | --- | --- | --- | --- |
| RSVPreF3+AS01 | Overall | 1,379,335 | <11 | 2.75 (0.46, 5.04) | 1.00 (0.17, 1.83) |
| RSVPreF | Overall | 682,267 | 13 | **6.91 (1.85, 11.97)** | 2.51 (0.67, 4.34) |

^a^ Statistically significant results are highlighted in bold

CI = Confidence Interval, GBS = Guillain-Barré Syndrome, IRR = Incidence Rate Ratio, PPV= Positive Predictive Value, RSV = Respiratory Syncytial Virus

Cell sizes <11 are suppressed to protect patient confidentiality

**eFigure 1: Hypothetical Example of Observation Period, Risk and Control Interval for Individual with a Qualifying GBS Outcome Following RSV Vaccination**

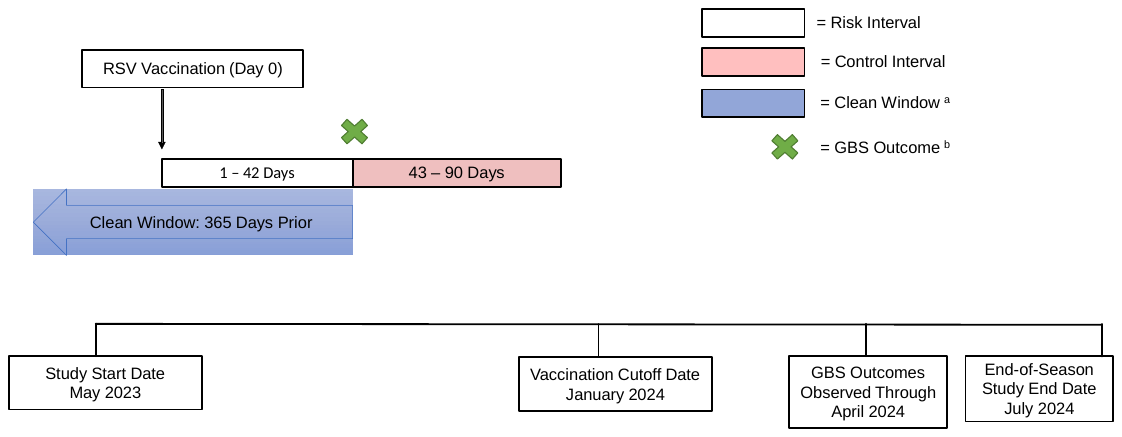

^a^ The clean window is relative to the outcome date; Risk and Control Intervals are relative to the vaccination date

^b^ Incident GBS outcomes identified using ICD-10-CM diagnosis code G61.0 in the primary diagnosis position on hospital inpatient claims

GBS = Guillain-Barré Syndrome, RSV = Respiratory Syncytial Virus

**eFigure 2: Secondary Analyses: IRR with Corresponding 95% CIs of GBS following RSV Vaccination for All Claims-Identified GBS Cases**

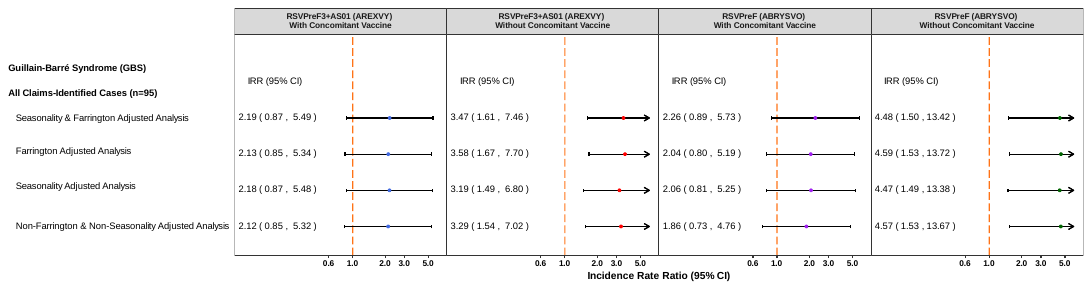

CI= Confidence Interval, GBS = Guillain-Barré Syndrome, IRR= Incidence Rate Ratio

**eFigure 3: Secondary Analyses: IRR with Corresponding 95% CIs of GBS following RSV Vaccination for PPV-Based Imputation Analyses and Chart-Confirmed GBS Cases**

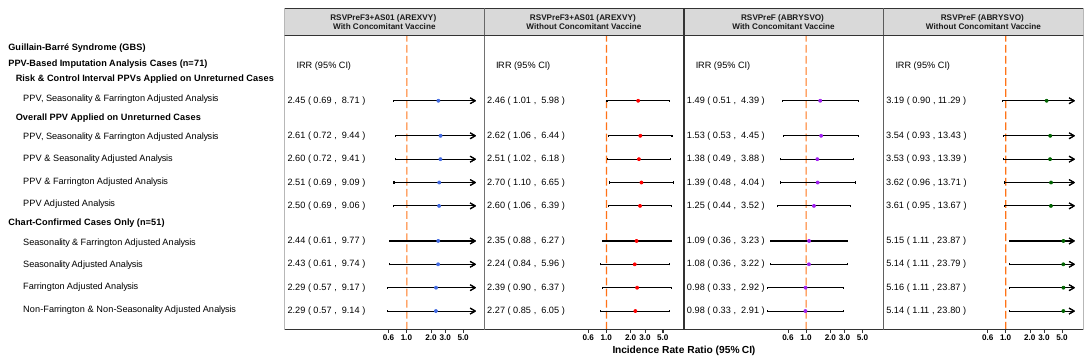

CI= Confidence Interval, GBS = Guillain-Barré Syndrome, IRR= Incidence Rate Ratio, PPV = Positive Predictive Value

**eFigure 4: Sensitivity Analyses: IRR with Corresponding 95% CIs of GBS following RSV Vaccination for PPV-Based Imputation Analyses, Chart-Confirmed, and All Claims-Identified GBS Cases**

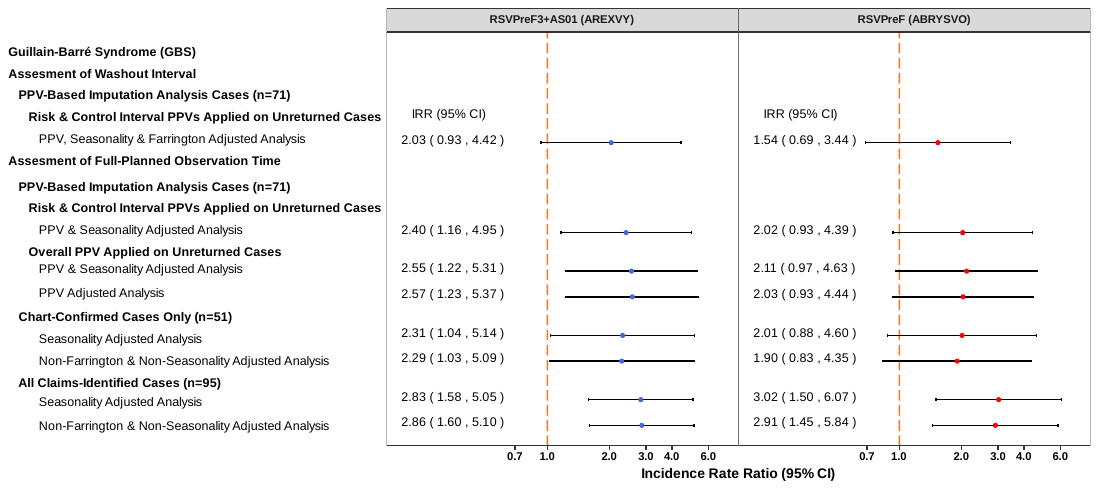

CI= Confidence Interval, GBS = Guillain-Barré Syndrome, IRR= Incidence Rate Ratio, PPV= Positive Predictive Value

**eFigure 5: Early-Season SCCS Analyses: IRR with Corresponding 95% CIs of GBS following RSV Vaccination for All Claims-Identified GBS Cases**

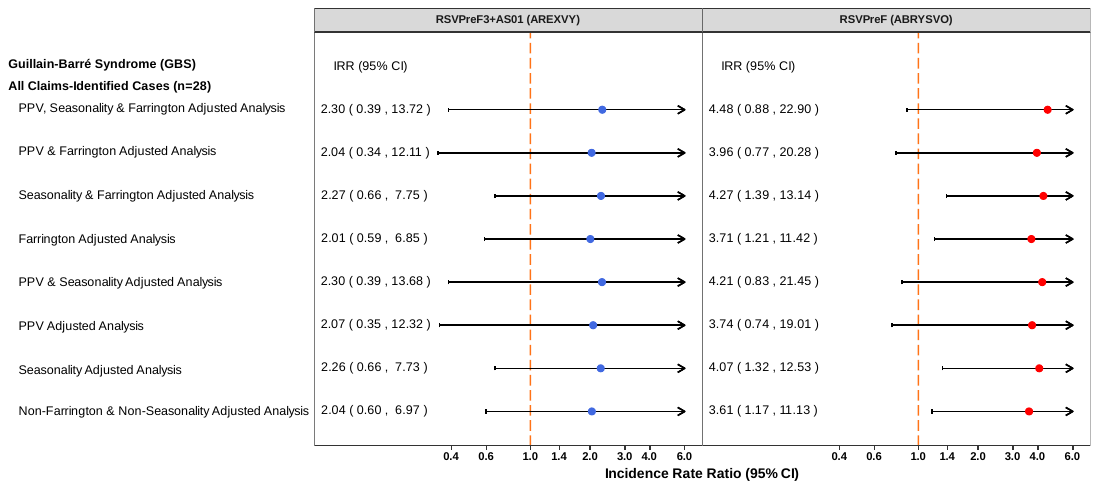

CI= Confidence Interval, IRR= Incidence Rate Ratio, PPV= Positive Predictive Value
